# Impact of Scabies on Quality of Life and Recent Advances in Management: A Systematic Review

**DOI:** 10.1101/2024.11.25.24317899

**Authors:** Sakshi Kumari, Swathi Srinivas, Zainab Siddiqua, Muhammad Saeed Qazi, Mahin Sheikh, Shree Rath, Maham Khan, Muhammad Daim Jawaid, Nayanika Tummala, Madho Mal

## Abstract

**Introduction:** Scabies are a highly transmittable disease of the skin. It is caused by a specific mite named Sarcoptes scabiei. Individuals who are infected with scabies have a miserable life, and their quality of life is poor. It impacts the quality of life by causing physical discomfort due to itching. Not only does it lead to the mental distress of a diseased individual, but it also leads to social isolation due to the stigma associated with this condition. This systematic review highlights the profound effect of scabies on the lives of affected individuals and the recent breakthroughs in the management of scabies.

**Purpose:** The purpose of this article is to shed light on the impact of scabies on the quality of life. We also discussed the advancements in the management of this disease, which impacts life physically and socially.

**Method:** We included randomized controlled trials, clinical trials, and observational studies. Studies with full-text access were included. Case reports, letters, and systematic/narrative reviews were excluded from the search. 180 records were screened up to March 2024, of which 129 articles were from PubMed and 51 from Google Scholar. 22 articles were thoroughly screened by two different reviewers independently and included in the review. Studies were carried out with no restrictions on age or sex, including the pediatric and adult populations.

**Conclusion:** Findings indicate a significant correlation between scabies and poor quality of life. Scabies most commonly affect children of lower socioeconomic status and in resource-limited settings. Scabies can lead to severe complications such as rheumatic heart disease, kidney diseases, and a variety of secondary bacterial infections; therefore, advances in treatment strategies are needed. Currently, permethrin and ivermectin have been used for treatment. Moxidectin, a new drug, is undergoing clinical trials to replace permethrin in regions with high resistance. The interventions need to be patient-centered, and new drugs need to be developed that have anti-parasitic actions and also lead to the prevention of secondary bacterial infections.

## Introduction

The infectious disease scabies is brought on by an infestation of the obligatory human parasite, Sarcoptes scabiei. Skin-to-skin contact, including intercourse, or, less frequently, contact with contaminated fabrics (such as clothes and towels), can result in the infestation. Less than 5 to 15 mites per host are present in classical scabies. Crusted scabies are typified by an extremely elevated mite burden in the afflicted person (1). It is typically seen occasionally as an institutional outbreak in hospitals, schools, retirement communities, and long-term care facilities in industrialized countries. Its incidence in underdeveloped urban and rural areas may approach 10% in the overall population and 60% in children (2). Around 300 million cases of scabies are reported annually, with a significant concentration of infections in Southeast Asia. Scabies is the sixth most prevalent skin condition in some locations, and summertime is when most instances are reported. (3)

Scabies were found to be the cause of 0.21% of disability-adjusted life years globally across all conditions examined by the Global Burden of Disease 2015 study. As a result, the degree of clinical pathology and the psychological effects of the disease may both have an impact on scabies morbidity. Furthermore, because of the disease’s rising incidence and resistance to therapy, scabies may worsen patients’ quality of life (QoL). QoL is affected by scabies to a moderate-to-severe degree. Depressive and anxiety scores are positively correlated with impaired QoL. (4) Nevertheless, Prompt management of scabies can avert sequelae like acute rheumatic fever and acute post-streptococcal glomerulonephritis, as well as secondary pyoderma. Scabies significantly lower a patient’s quality of life. (3)

A comparison study conducted in Fiji showed that ivermectin outperforms permethrin. When azithromycin and ivermectin were taken together, however, the incidence of impetigo and scabies decreased just as much as when ivermectin was taken alone. After mass medication administration, the percentage of impetigo lesions containing pyogenic streptococci decreased. After the azithromycin mass drug administration, there was a brief rise in the percentage of S. aureus bacteria that were resistant to macrolides. (5)

The typical method for diagnosing scabies involves identifying the mite, eggs, or excrement through microscopic analysis of scales that are removed from the skin by scraping. The adhesive tape test and the burrow ink test are two further diagnostic techniques. Some sophisticated non-invasive methods, including optical coherence tomography, dermatoscopy, in vivo reflectance confocal microscopy, and video dermatoscopy, have shown increased efficacy in the diagnosis of scabies in recent years (2).

A study shows At the 4-week mark, Ivermectin had a cure rate of 78.5% with a single dose, whereas 59.5% of patients responded well to a single treatment of 10% sulfur ointment. At the 2-week follow-up, a single ivermectin dose was just as effective as a single application of sulfur 10% ointment. At the 4-week follow-up, ivermectin was better than sulfur 10% ointment after the treatment was repeated. Ivermectin’s delayed clinical response raises the possibility that it is ineffective against all phases of the parasite’s life cycle. Another study shows that at the 2-week follow-up, two treatments of ivermectin were just as effective as two applications of permethrin 2.5% cream. At the 4-week follow-up, ivermectin was just as effective as permethrin 2.5% cream after the treatment was repeated. (6,7)

Although permethrin has a higher treatment cost per case, it can offer a considerable efficacy of 88% and acceptance in 100% of instances. Ivermectin is less expensive and has efficacy and acceptance rates of 84% and 96%, respectively. Benzyl benzoate was the least expensive medication and delivered 80% for both rates. The least effective and most costly product was sulfur ointment. (8)

This comprehensive systematic review emphasizes the significant impact scabies have on afflicted people’s lives as well as the most current advancements in scabies treatment. This article’s goal is to evaluate how scabies affect quality of life and discuss recent advancements in their management. We also examined its nature, its impact on well-being, optimal medications, their effectiveness, and strategies to decrease its prevalence.

### Objectives

To assess the impact of scabies on quality of life and recent advances in management

## Methodology

### Aims and Objectives

“What is the impact of scabies infection on the quality of life, and what are recent advancements made in the management of this disease?

### Search Strategy

PubMed was searched using a combination of keywords “Scabies, Quality of life/Psychology and Management”, and included interventions to identify planned, ongoing, terminated, and completed RCTs and Clinical trials. Full-text publications were retrieved from 2014 - 2024. If no full-text publications were found Google Scholar was also searched to retrieve full texts. Following is the full PubMed search strategy, which is modified for the Cochrane Database.

### Scabies and QOL without Cochrane trial filter

*Scabies [tw] OR “Sarcoptes scabiei” [tw] OR “Mange, Sarcoptic” [tw] OR “Sarcoptic Mange” [tw] OR Scabies [mh] AND (“Quality of life” [tw] OR Stigma [tw] OR “Psychological wellbeing” [tw] OR “Patient satisfaction” [tw] OR “Psychological resilience” [tw] OR “Life Quality” [tw] OR “Health-Related Quality Of Life” [tw] OR “Health-Related Quality Of Life” [tw] OR HRQOL OR “Life Styles”[tw] OR Lifestyle* [tw] OR “Lifestyle Induced Illness” [tw] OR “Lifestyle Factors” [tw] OR “Factor, Lifestyle” [tw] OR “Lifestyle Factor” [tw] OR “Value of Life” [tw] OR “Quality of Life/psychology” [mh])*

### Scabies and Management with Cochrane Trial Filter

*((Scabies [tw] OR “Sarcoptes scabiei” [tw] OR “Mange, Sarcoptic” [tw] OR “Sarcoptic Mange” [tw] OR Scabies [mh]) AND (Treatment* [tw] OR Diagnos* [tw] OR Strateg* [tw] OR “Disease Management*” [tw] OR Manag* [tw] OR Advancement* [tw] OR “Management, Disease” [tw] OR “Disease Management” [mh])) AND (randomized controlled trial [pt] OR controlled clinical trial [pt] OR randomized [tiab] OR placebo [tiab] OR drug therapy [sh] OR randomly [tiab] OR trial [tiab] OR groups [tiab] OR animals [mh] NOT humans [mh])*

### Cochrane Central Search Strategy

#1 MeSH descriptor: [Scabies] explode all trees

#2 (Scabies): ti,ab,kw (Word variations have been searched)

#3 (Sarcoptes scabiei): ti,ab,kw (Word variations have been searched)

#4 (Mange, Sarcoptic): ti,ab,kw (Word variations have been searched)

#5 (Sarcoptic Mange): ti,ab,kw (Word variations have been searched)

#6 #1 OR #2 OR #3 OR #4 OR #5

#7 MeSH descriptor: [Disease Management] explode all trees

#8 (Treatment): ti,ab,kw (Word variations have been searched)

#9 (Diagnosis): ti,ab,kw (Word variations have been searched)

#10 (strategies): ti,ab,kw (Word variations have been searched)

#11 (advancement): ti,ab,kw (Word variations have been searched)

#12 #7 OR #8 OR #9 OR #10 OR #11

#13 #6 AND #12

#14 MeSH descriptor: [Quality of Life] explode all trees

#15 (Stigma): ti,ab,kw (Word variations have been searched)

#16 (Quality of life): ti,ab,kw (Word variations have been searched)

#17 (Psychological wellbeing): ti,ab,kw (Word variations have been searched)

#18 (Patient satisfaction): ti,ab,kw (Word variations have been searched)

#19 (Psychological resilience): ti,ab,kw (Word variations have been searched)

#20 (Life Quality): ti,ab,kw (Word variations have been searched)

#21 (Health-Related Quality of Life): ti,ab,kw (Word variations have been searched)

#22 (Value of Life): ti,ab,kw (Word variations have been searched)

#23 #14 OR #15 OR #16 OR #17 OR #18 OR #19 OR #20 OR #21 OR #22

#24 #6 AND #23

### Eligibility Criteria

1. RCT, Clinical Trials, and Observational studies are included
2. Studies conducted between 2014 - 2024.
3. Studies were carried out with no restrictions on age and sex including the pediatric and adult population.
4. Studies report recent advancements in the diagnosis, understanding, and treatment of scabies.
5. Studies exploring prevalence, risk factors, stigma, and physical symptoms associated with scabies.
6. Studies focusing on the impact of scabies infection on quality of life.
7. This review did not include case reports, letters to editors, reviews, and case series.
8. Articles in languages other than English are also excluded from the review.

### Screening and Selection

The bibliographic database was managed through Mendeley. Two independent reviewers initially assessed the paper based on its title and abstract. Case reports, letters, and systematic/narrative reviews were excluded from the search. Papers containing relevant keywords in their titles and abstracts were chosen for full-text review. If a paper lacked an abstract but its title related to the review’s objectives, it was also considered for full-text screening. Both reviewers undertook a detailed examination of the selected full-text papers. Those meeting all selection criteria were then subjected to data extraction. Additionally, the reviewers manually examined the reference lists of all selected articles to find any further relevant papers. Any disagreements between the reviewers were resolved through discussion.

### Data Extraction and Quality Assessment

Data extraction was done by one reviewer using a standardized form and was verified by another reviewer. The study was assessed based on study design, participants’ demographics, intervention, outcome measures, and safety profile. The quality assessment process was conducted by an independent reviewer and validated by a second reviewer through RevMan. For RCT, ROB2 was used. Disagreements for each criterion were settled through discussion.

One significant challenge during the data collection process was the unavailability of free full-text articles for many studies. This limitation hindered our access to comprehensive data, as we relied on complete texts to extract relevant information. Additionally, several studies were not available in English, which further restricted our access to crucial data. The language barrier prevented us from including potentially valuable studies in our analysis.

Another obstacle we faced was encountering studies with incomplete or insufficient data. Some studies lacked proper documentation of their methodology, making it difficult to assess the reliability and validity of their findings. Without a clear understanding of the study methodology, we were unable to incorporate these studies into our analysis with confidence.

Study findings included in the study are summarised below:

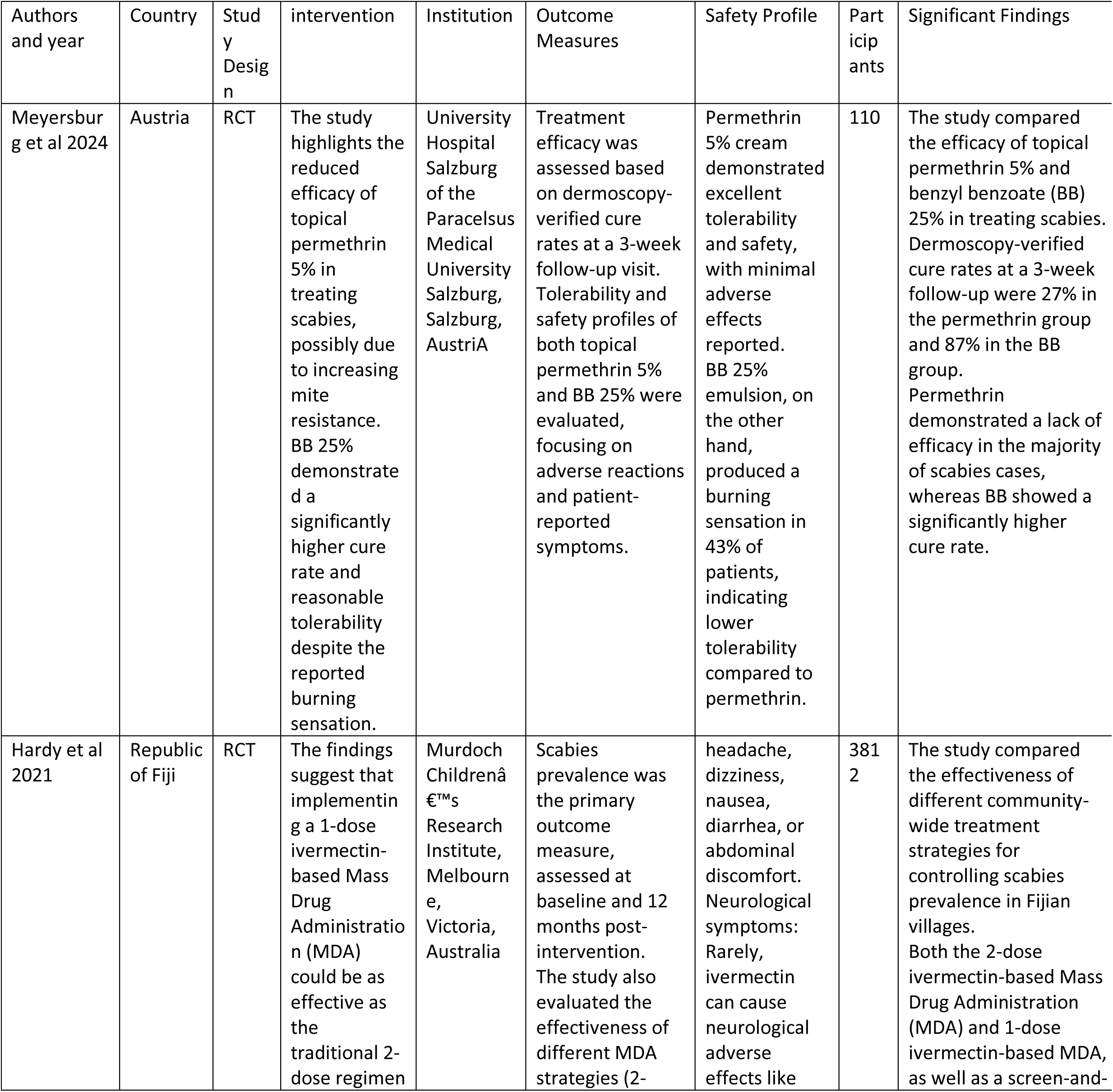
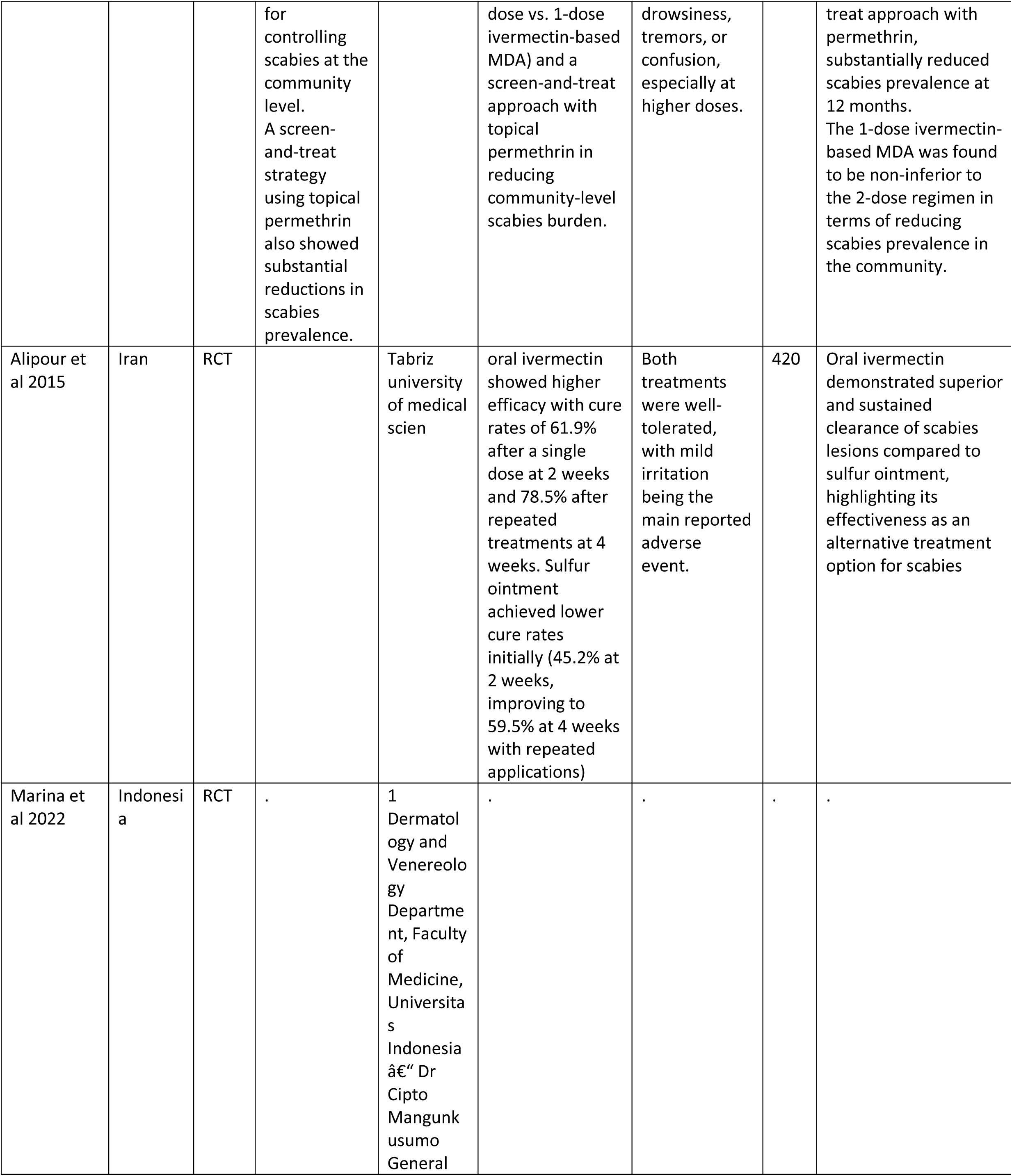
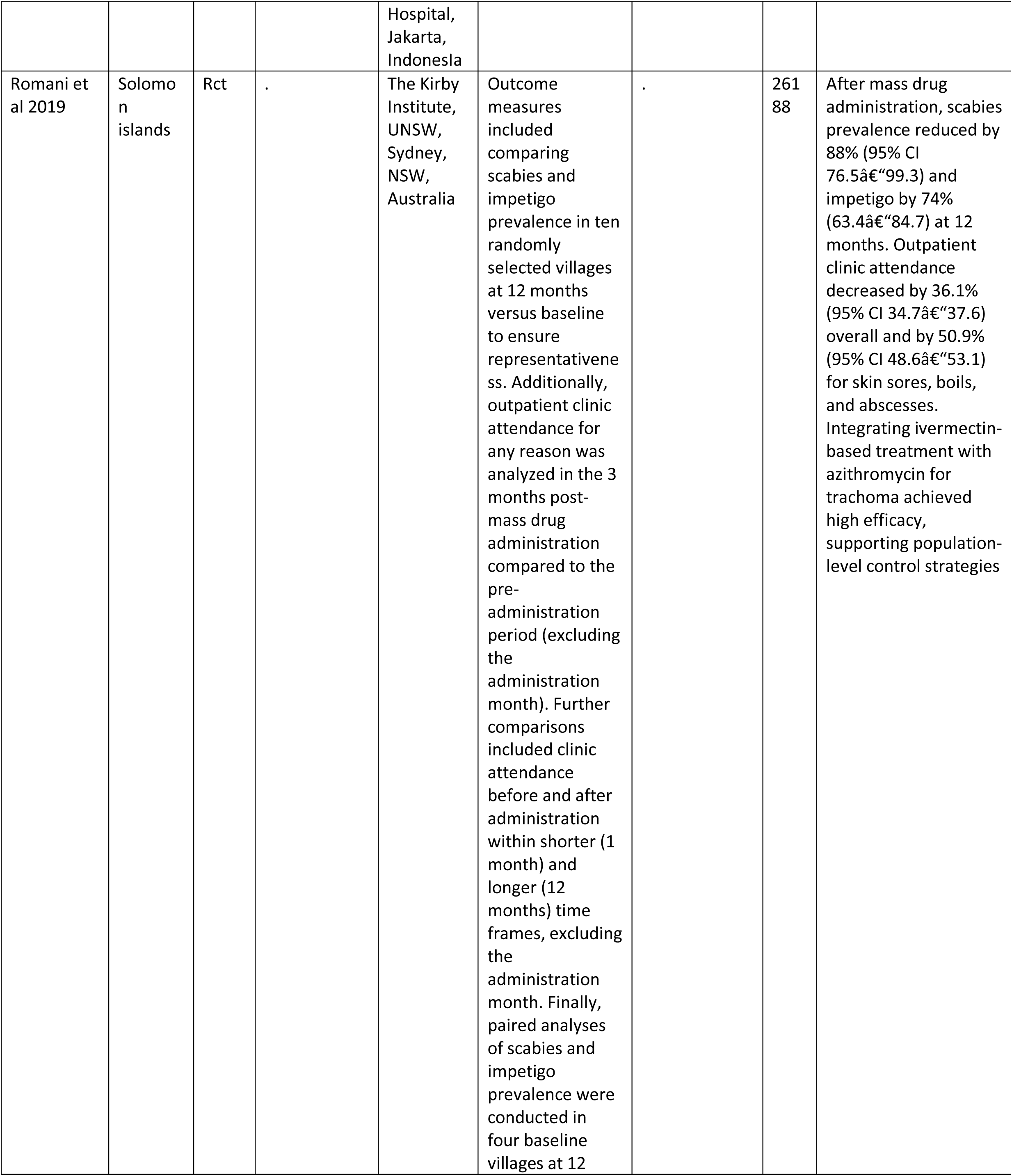
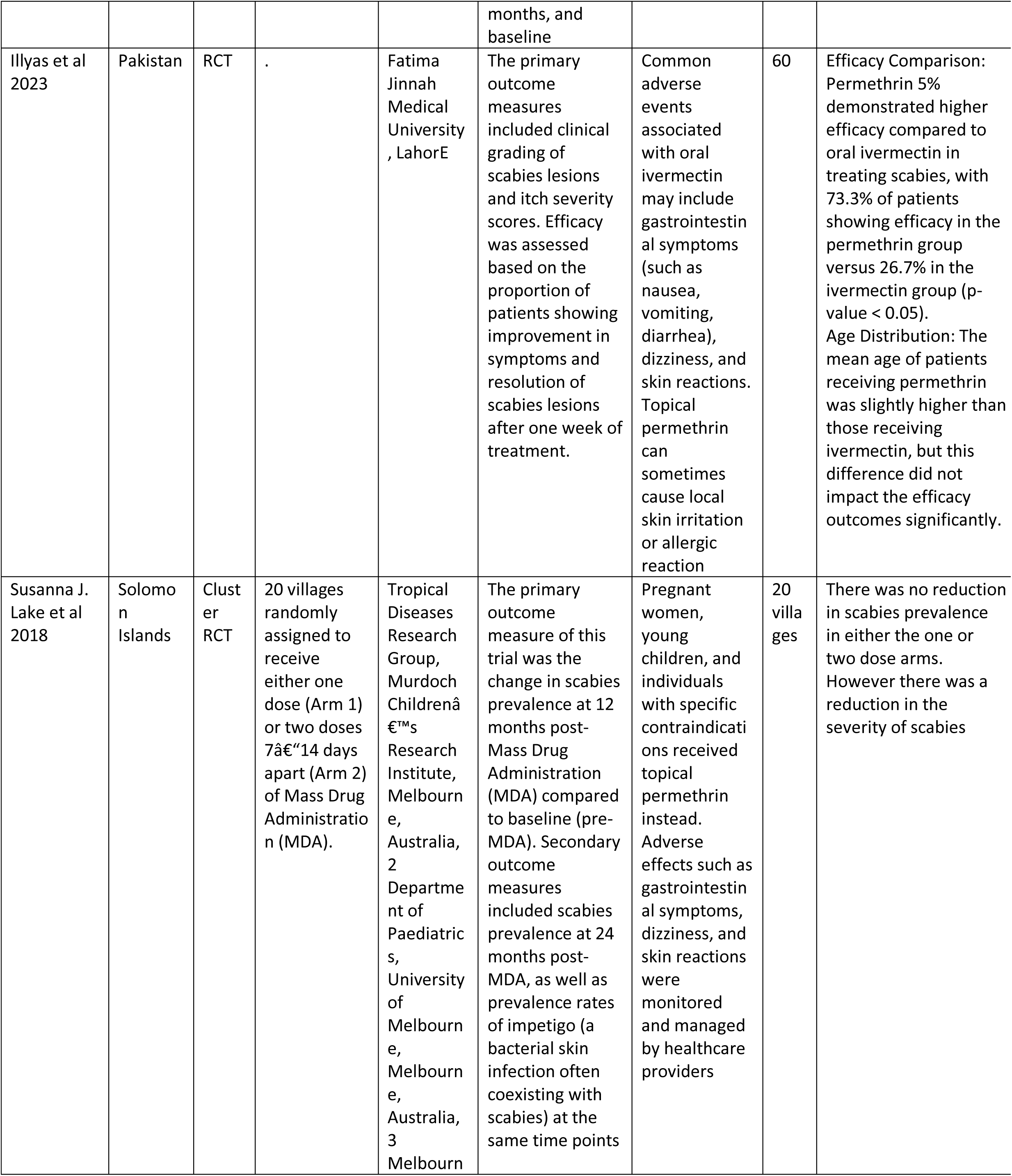
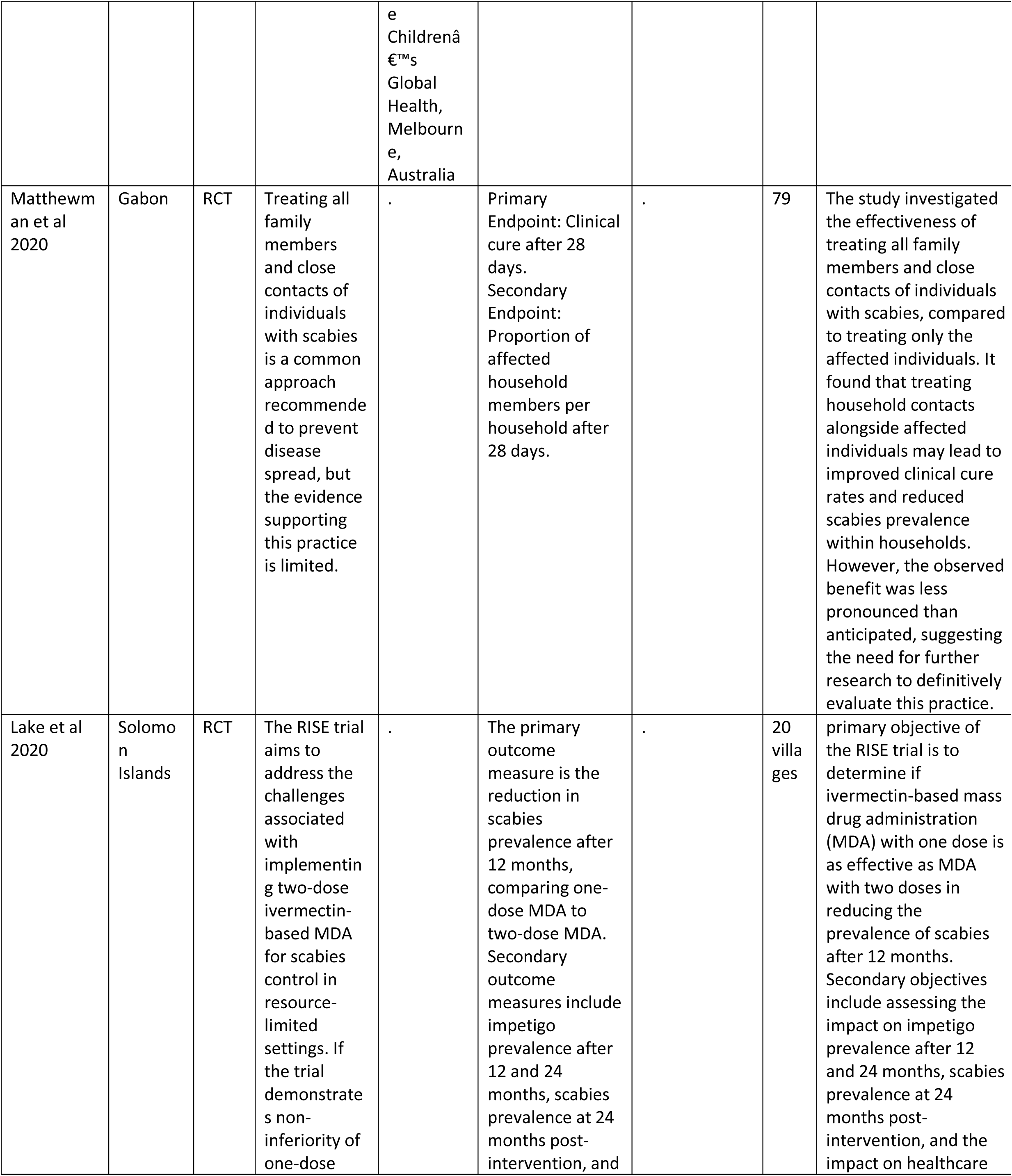
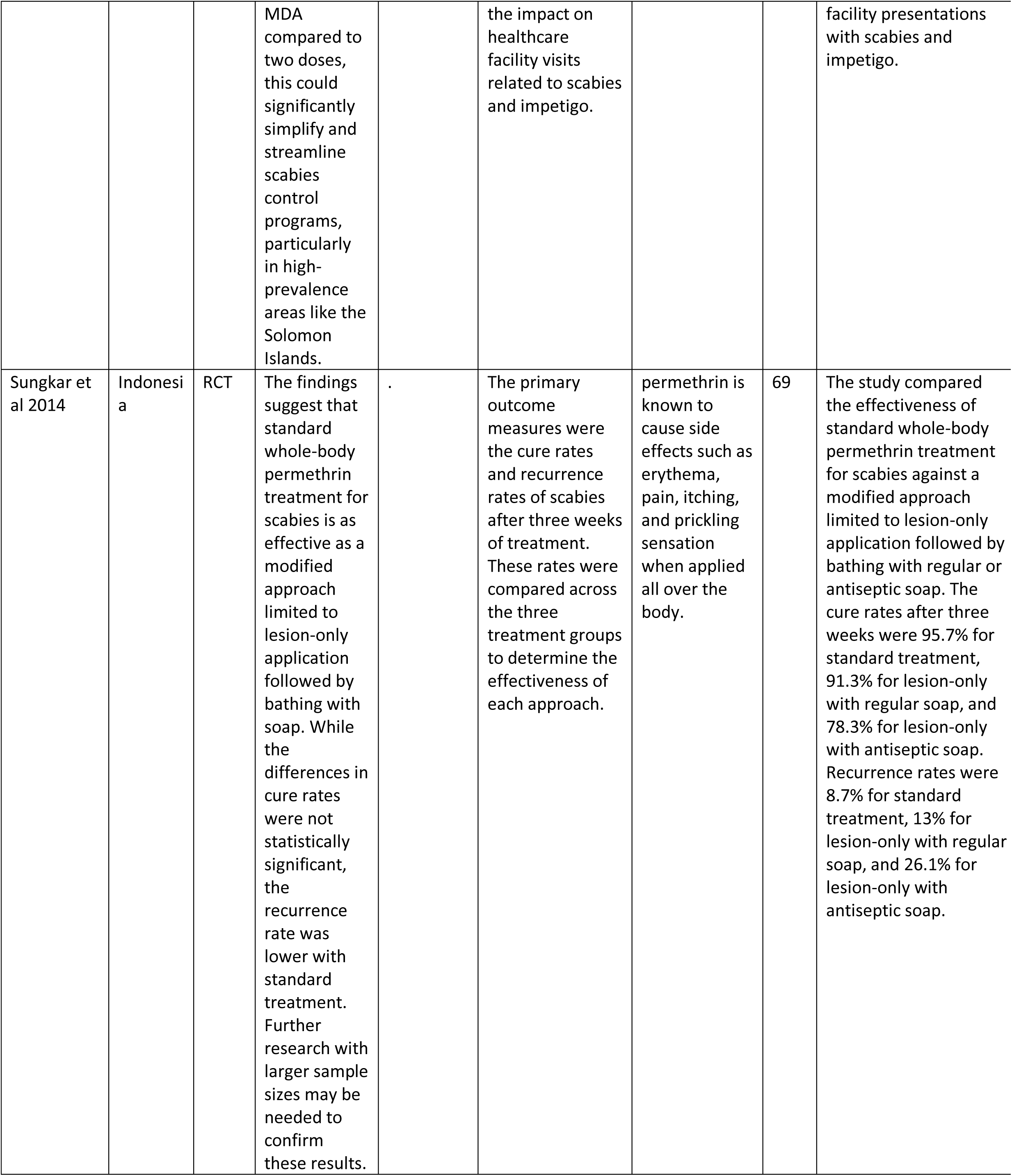
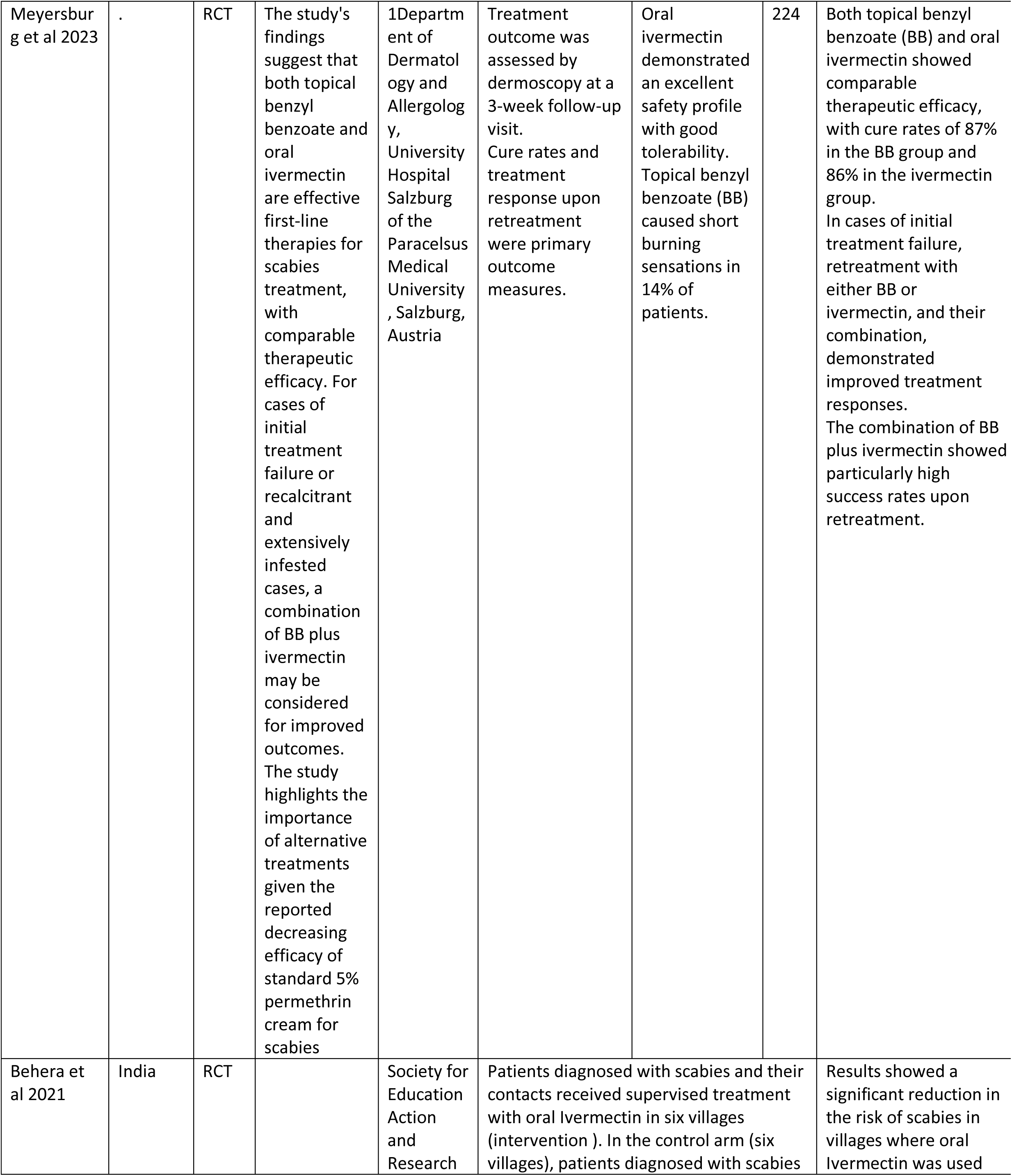
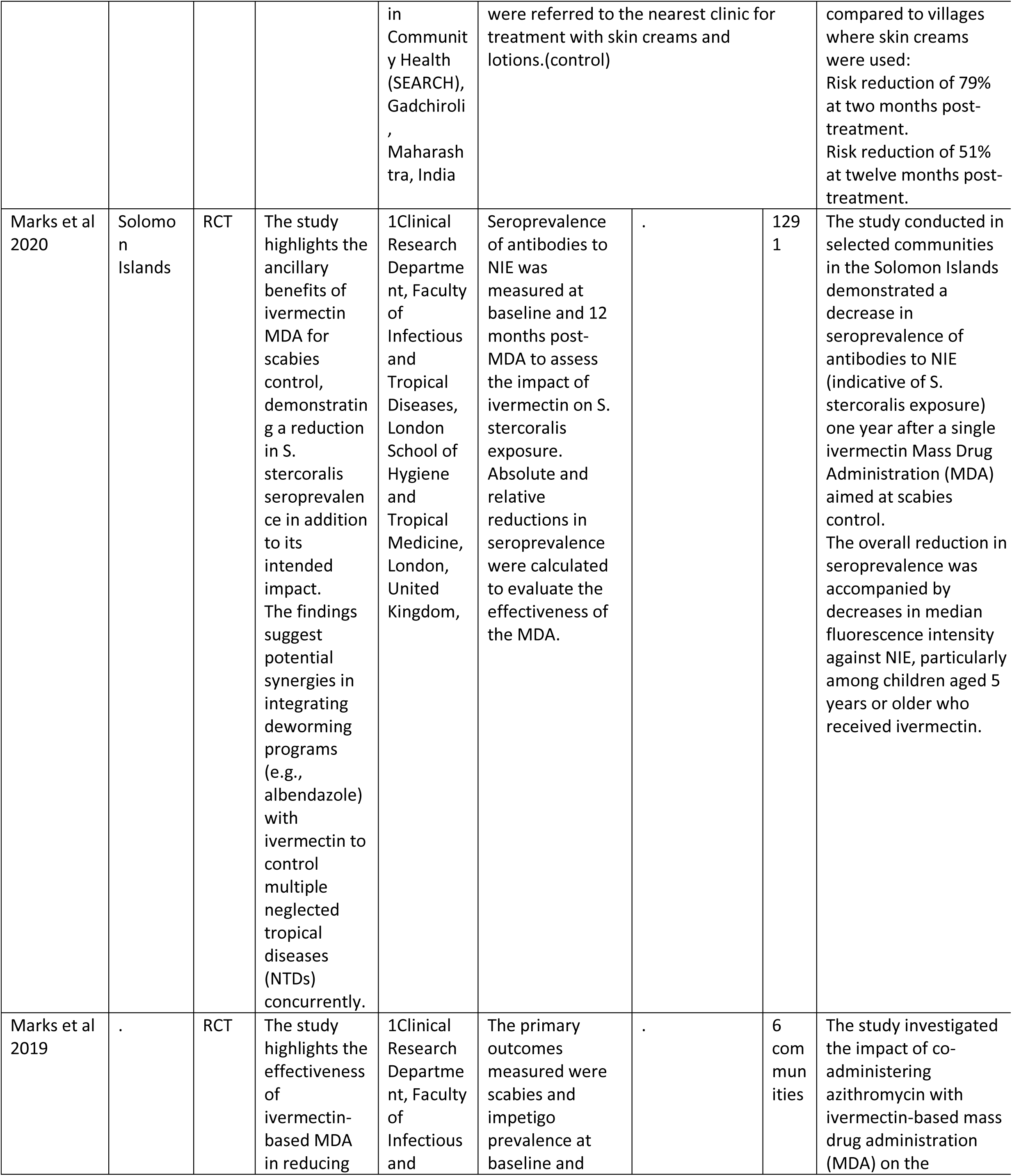
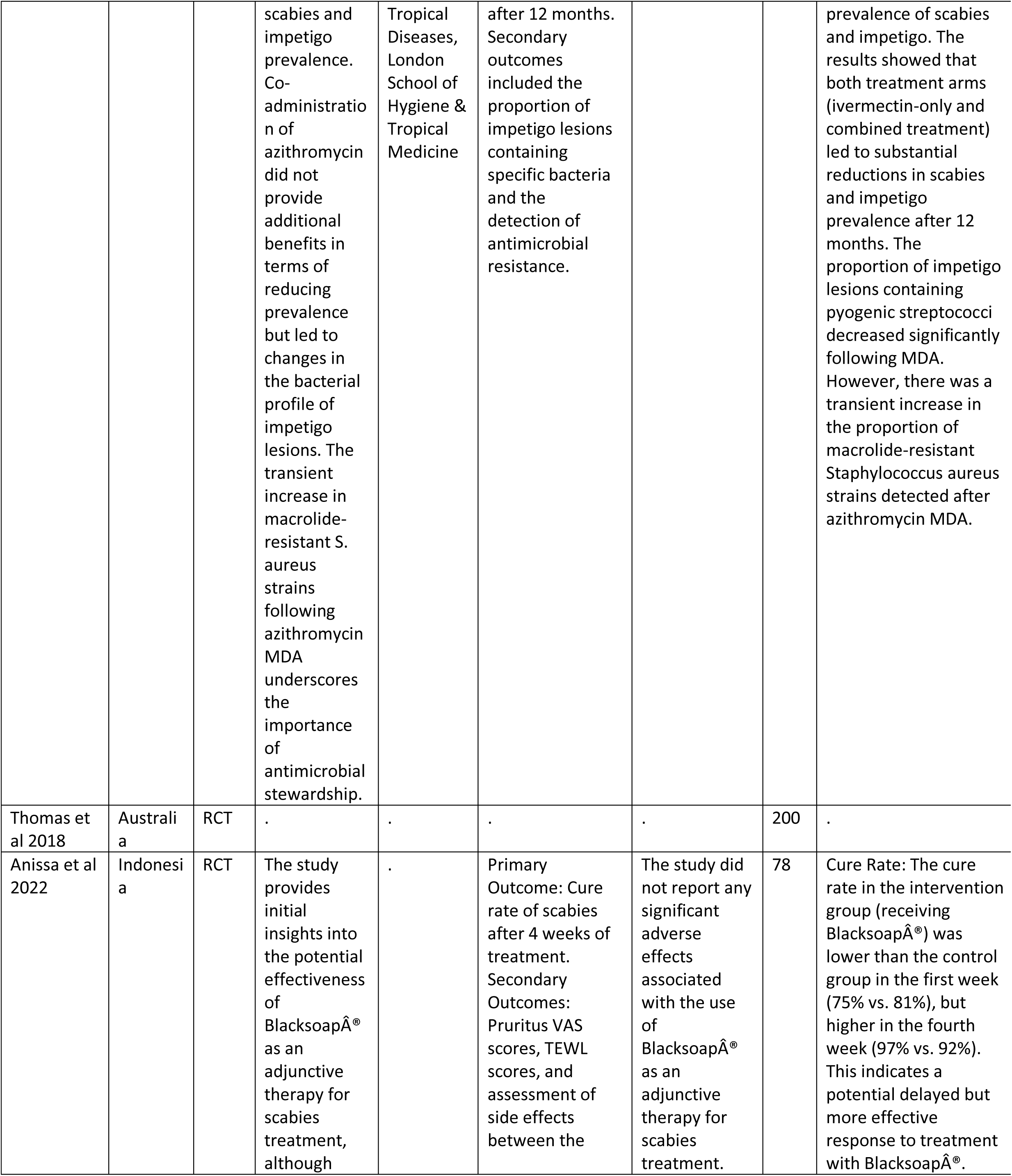
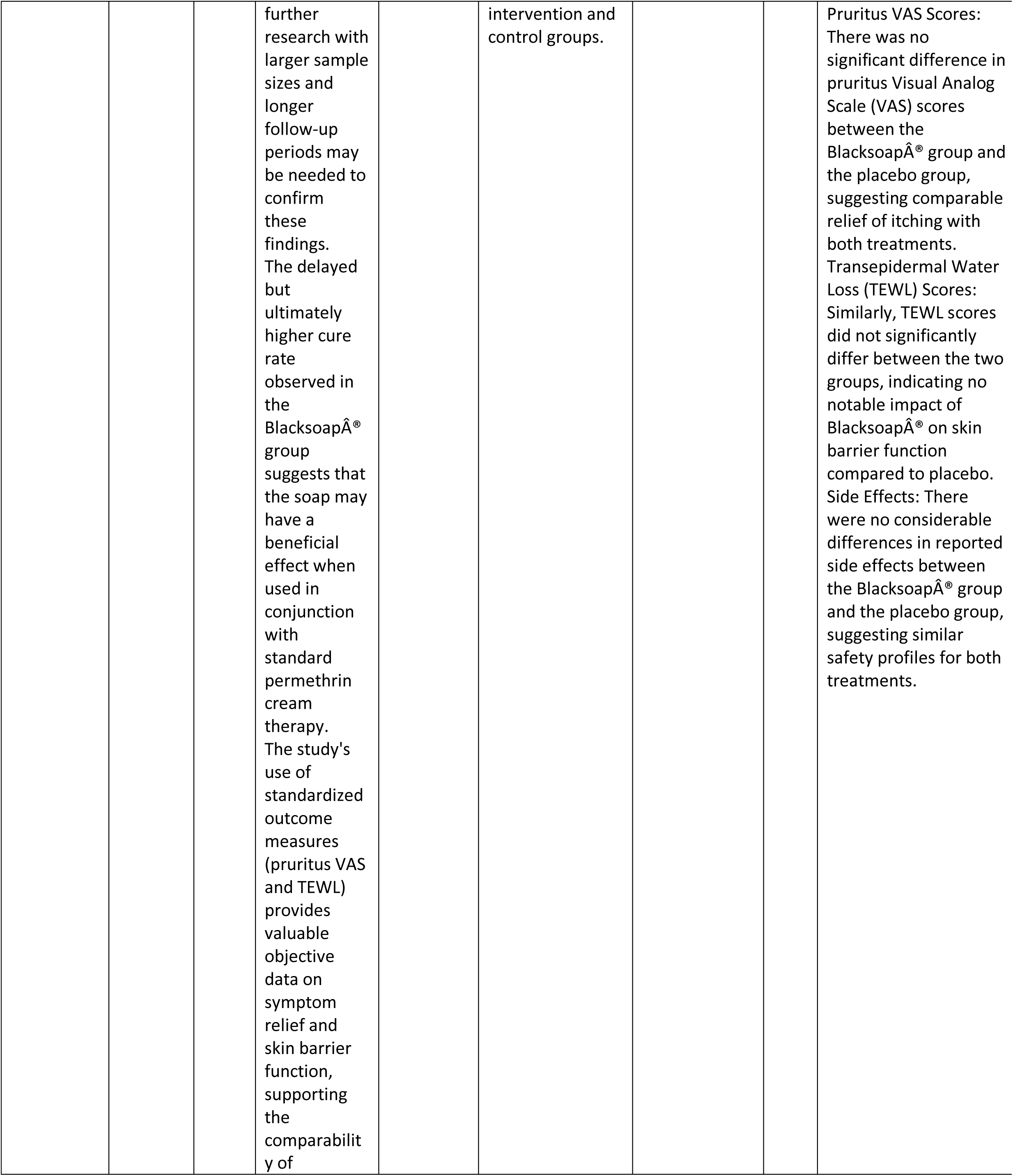
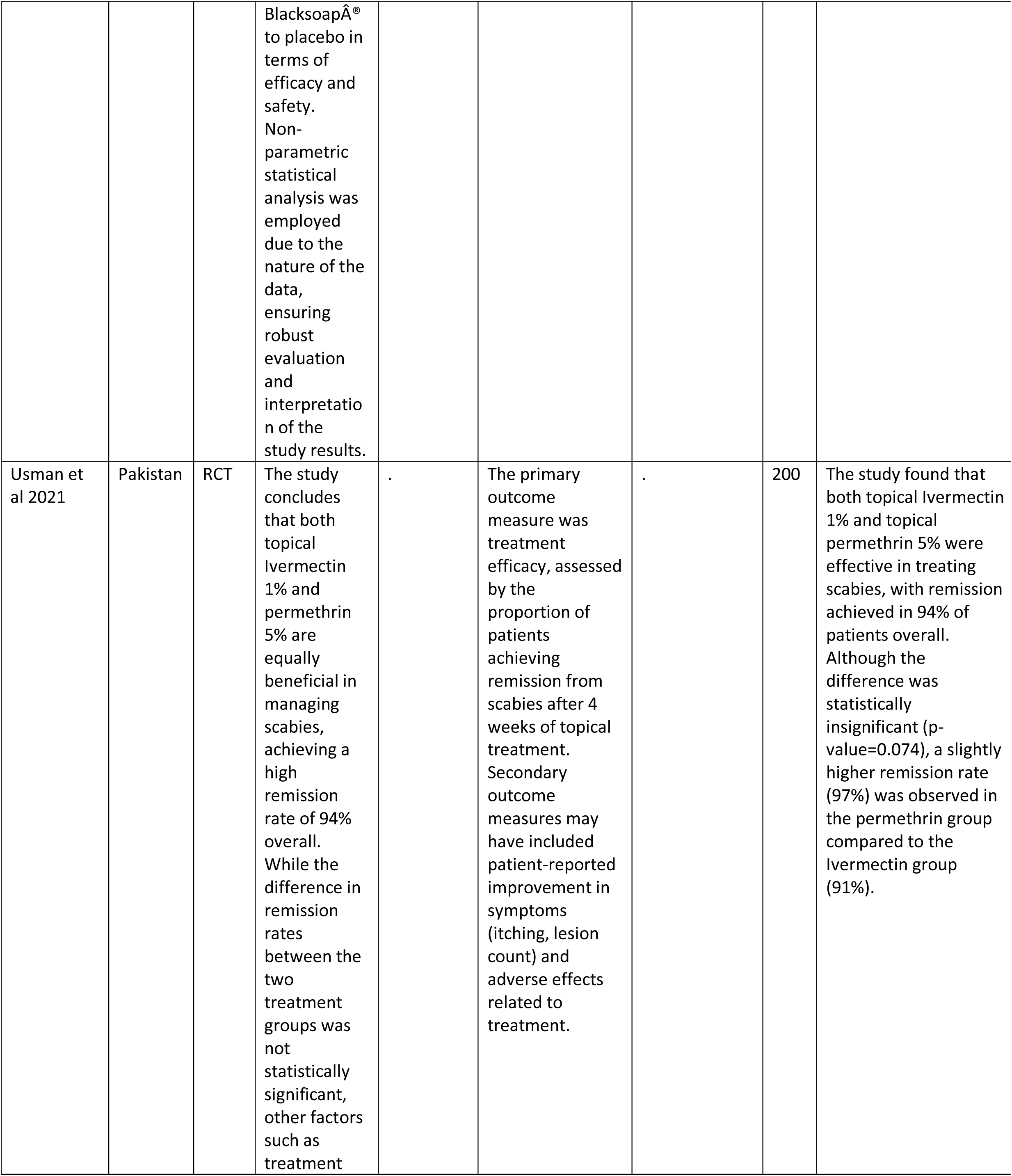
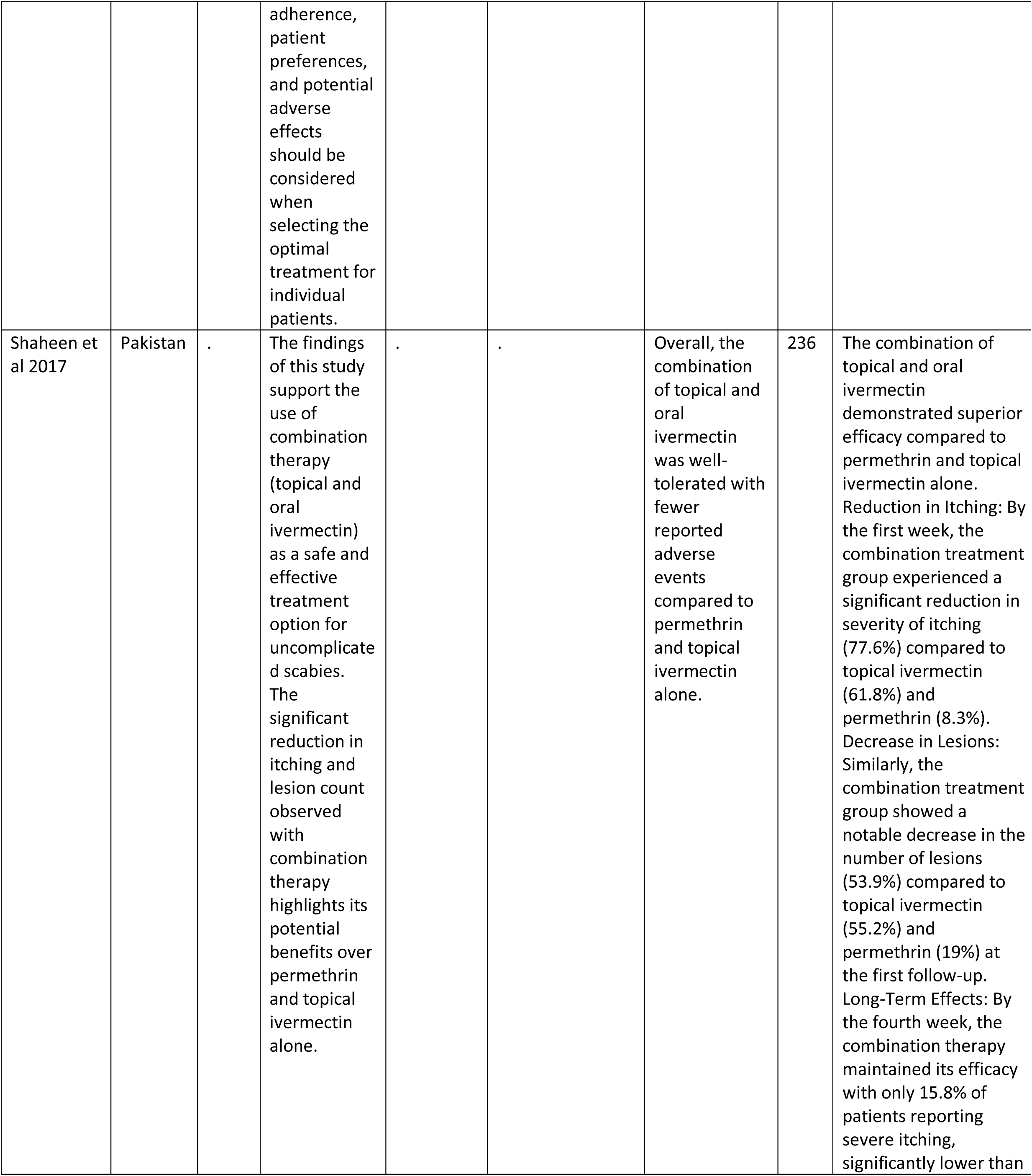
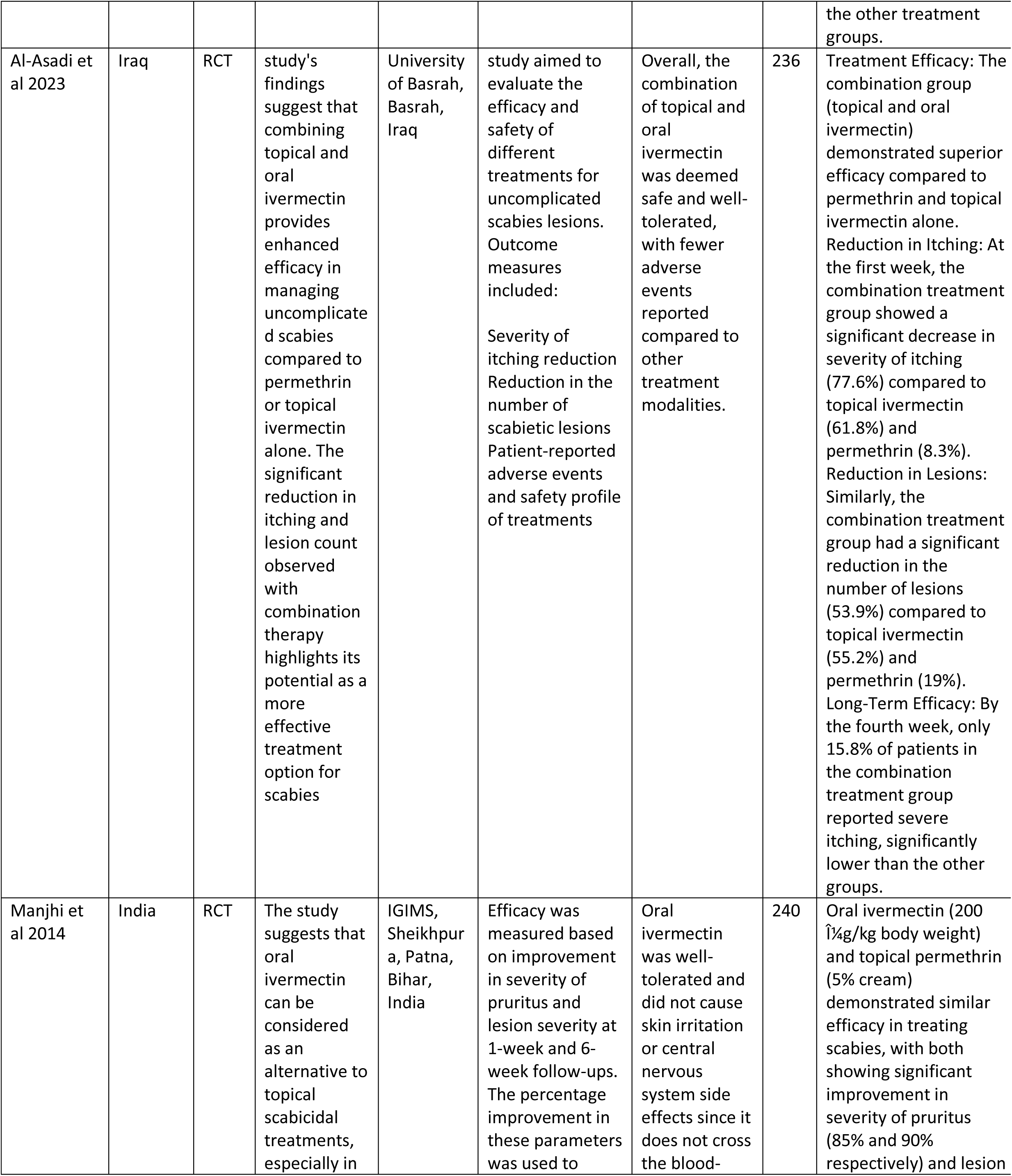
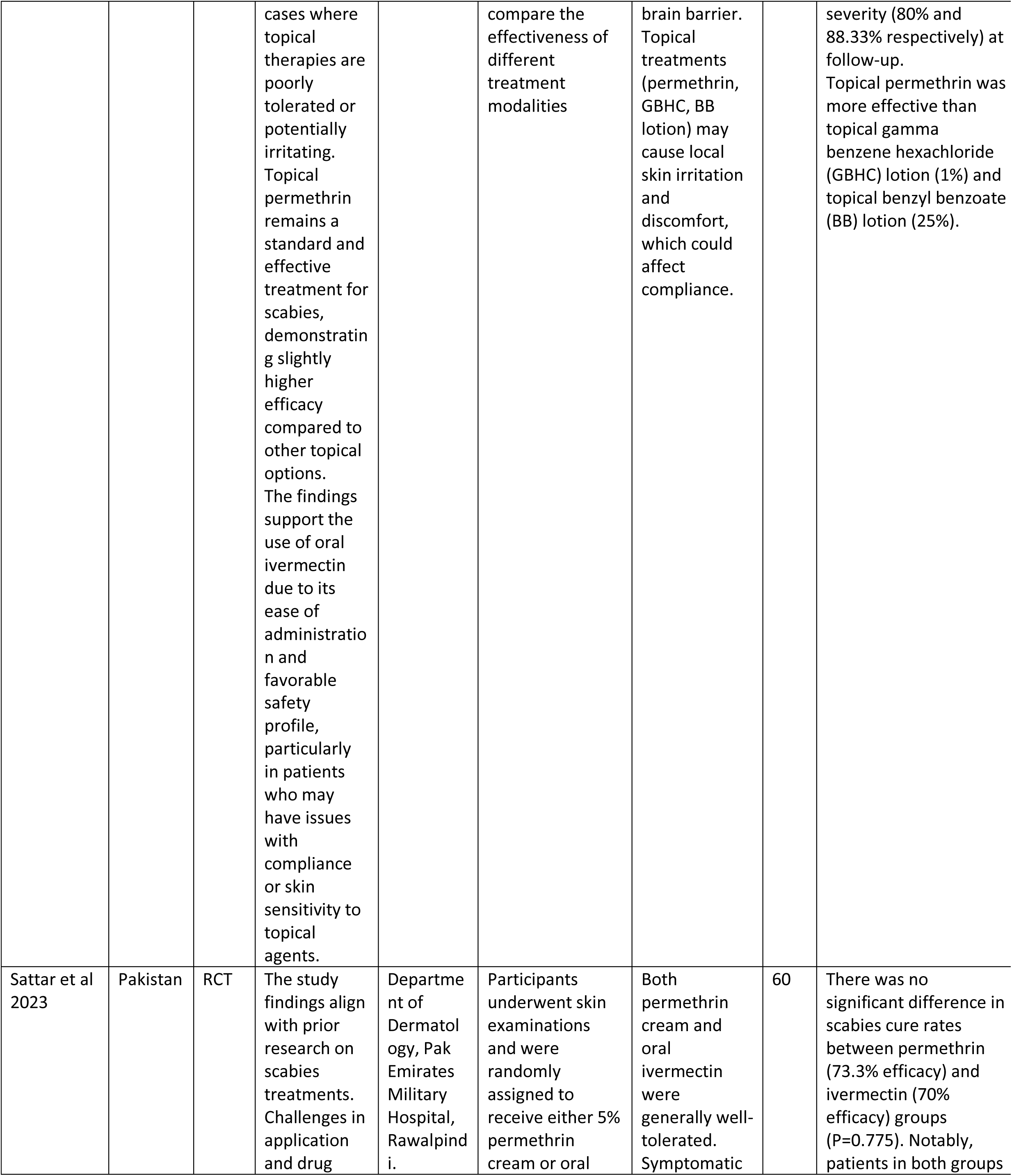
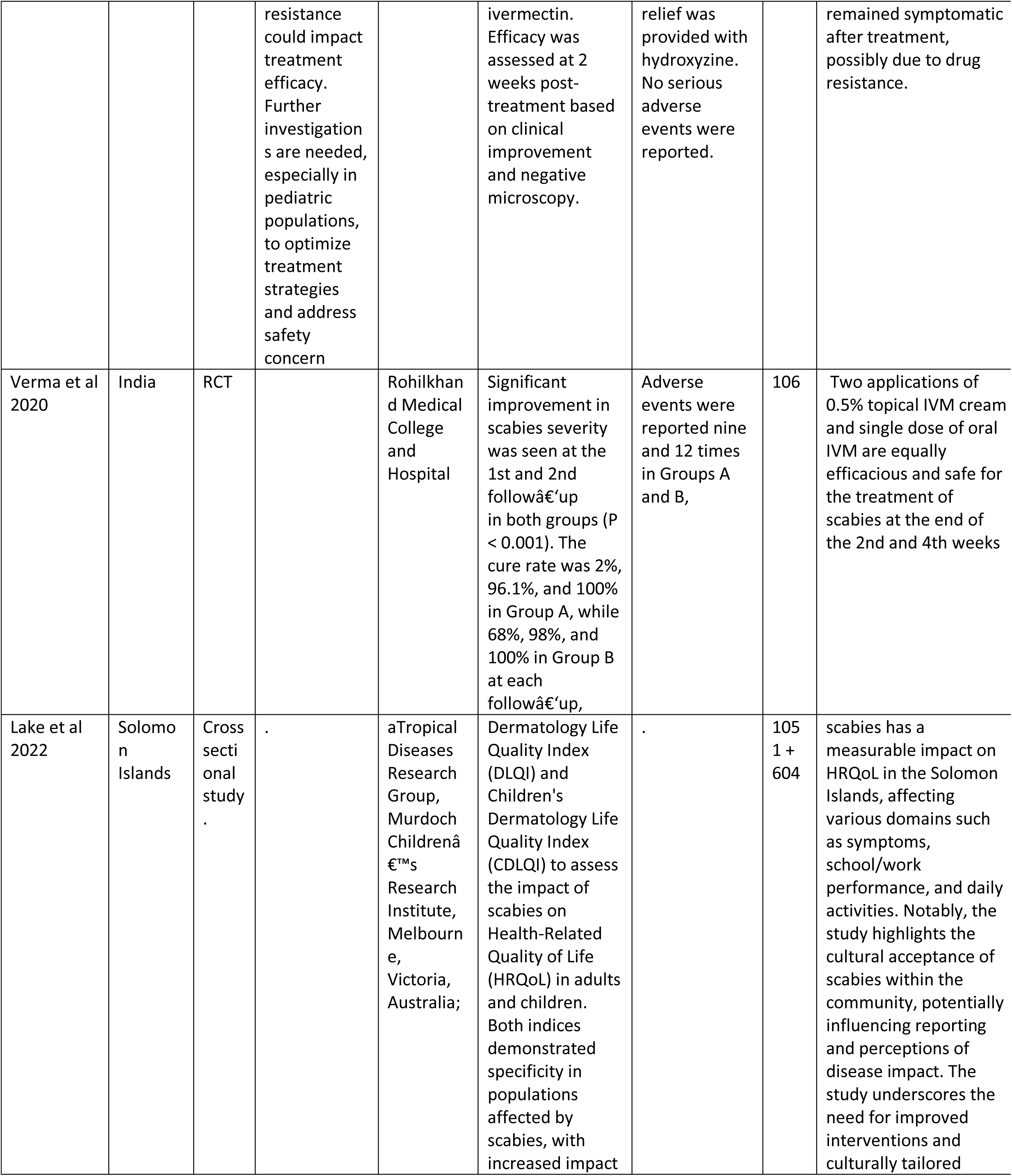
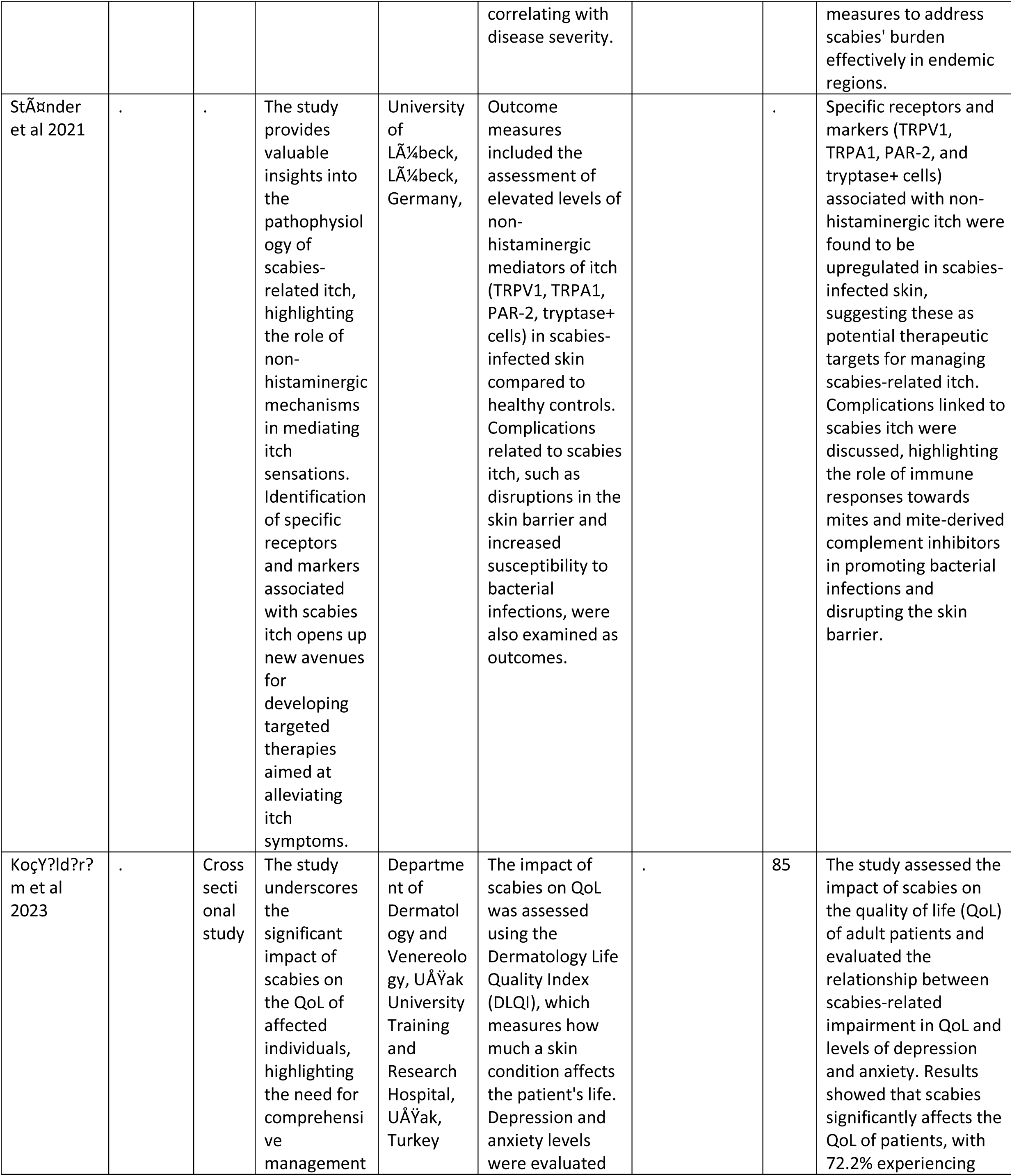
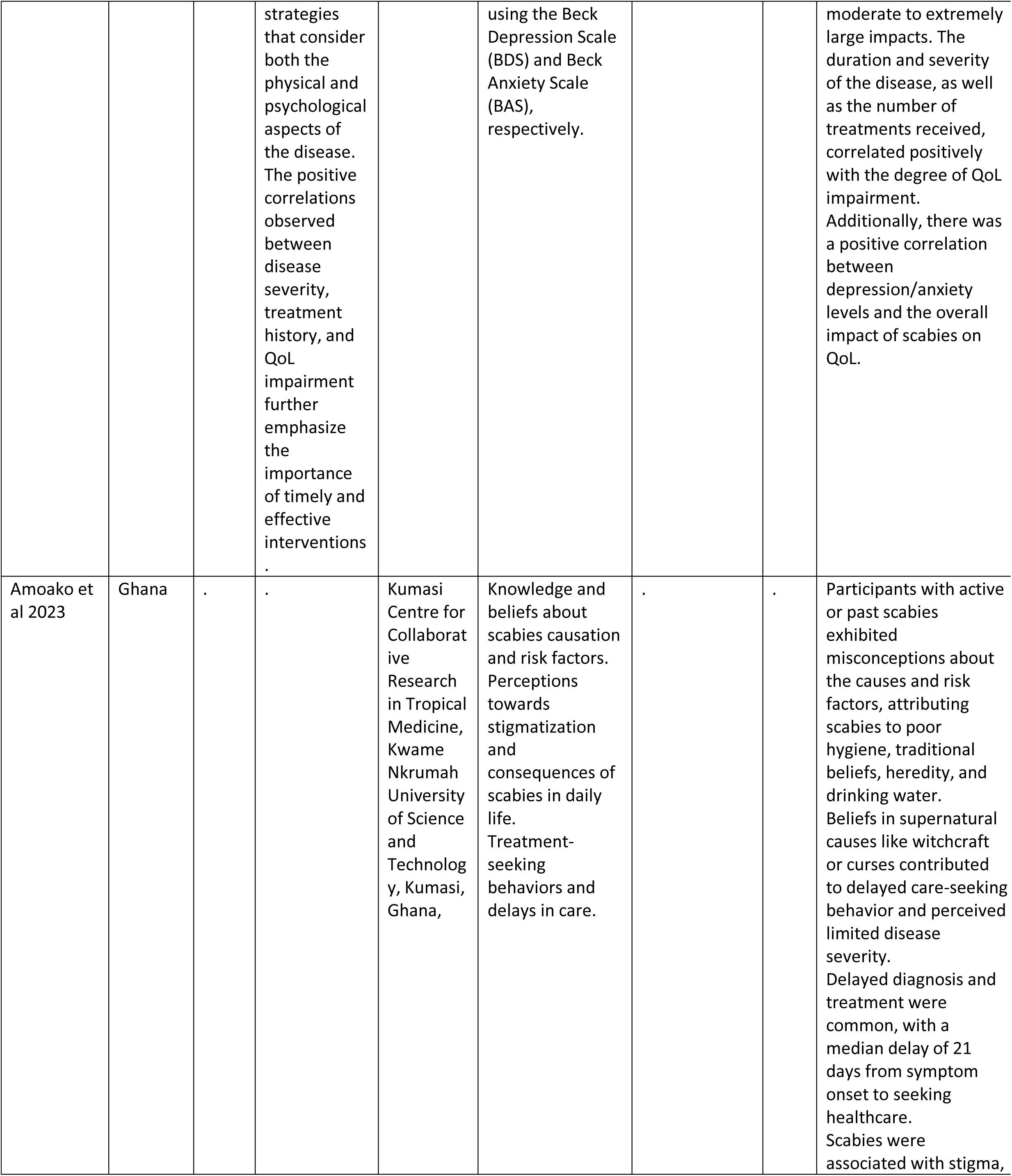
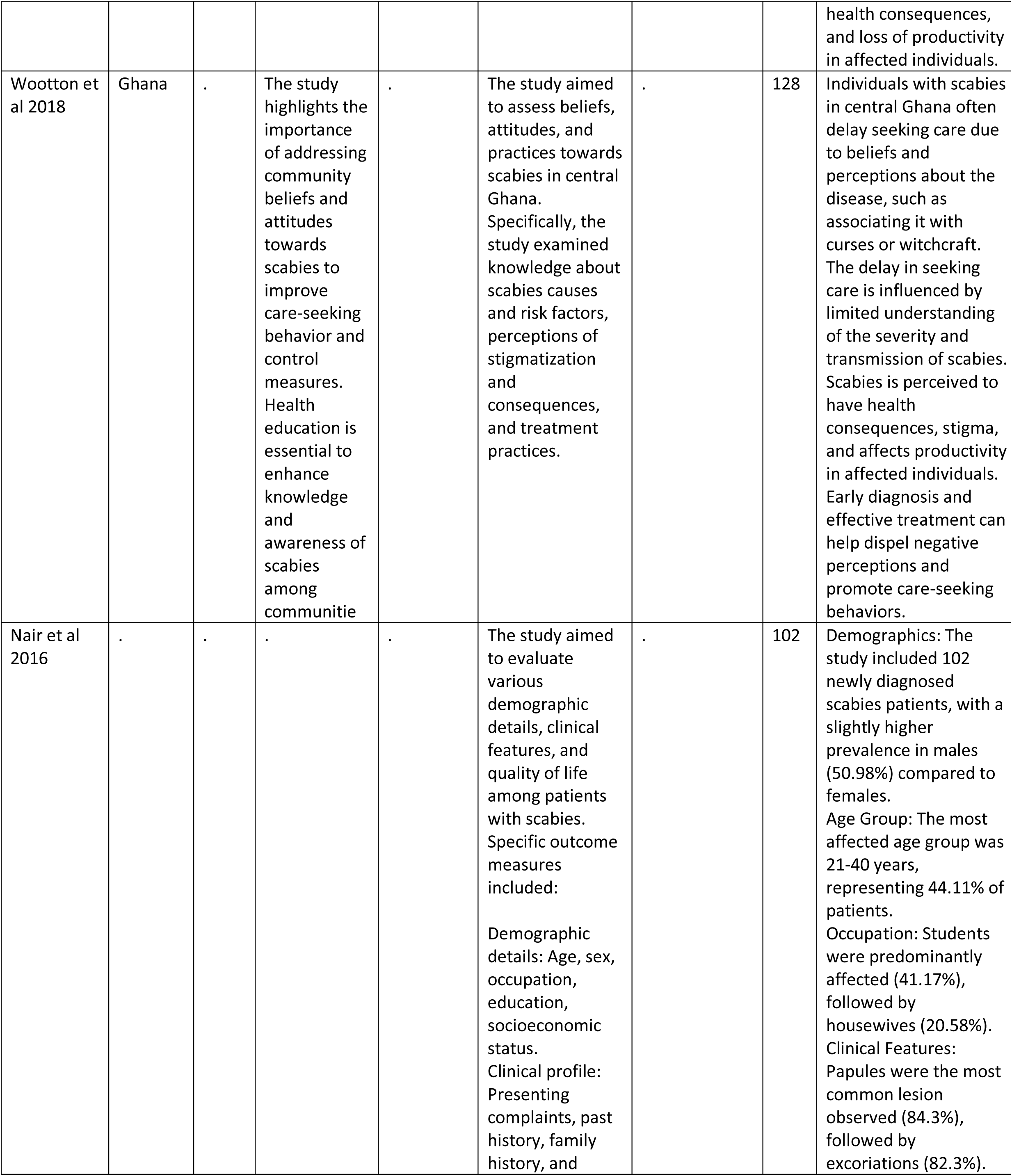
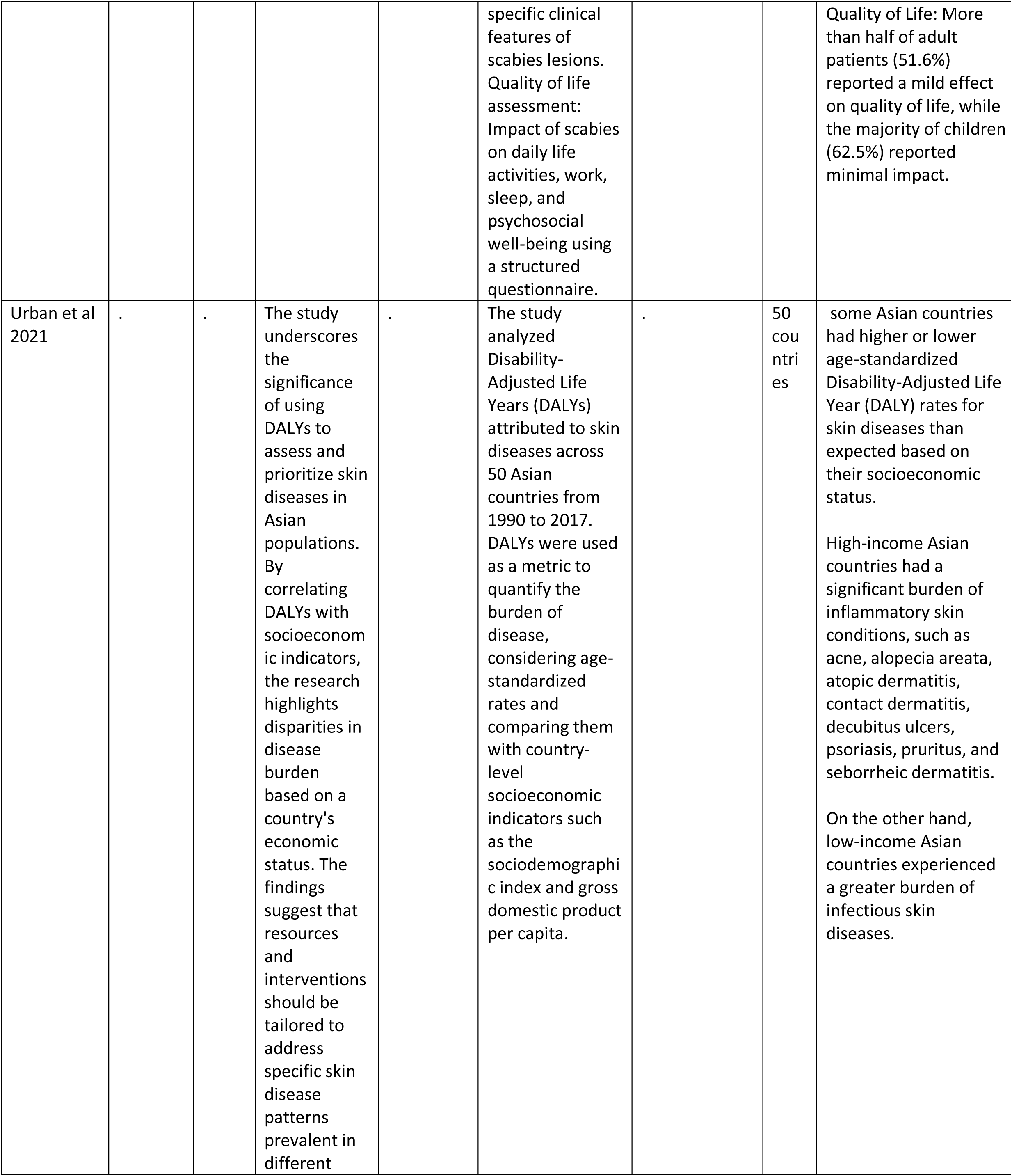
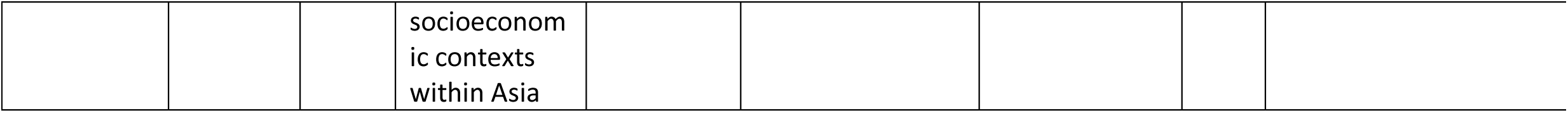

## Results

### 3.1. Comprehensive literature search

The selection procedure of all records received through the literature search is illustrated in Figure 1. A total of 242 records were screened, of which 168 articles were from Cochrane Central and 73 articles from PubMed. Of these, 16 records were removed after being found duplicated. 226 articles were screened for title and abstract and 96 articles were removed for not matching criteria. 130 reports were sought for retrieval and 70 of them were not retrieved due to lack of text. 61 articles were assessed for eligibility, out of which 31 articles were removed due to various reasons. Finally, 29 studies are included in this review. The Prisma Flow chart is given below:

**Figure 1:**
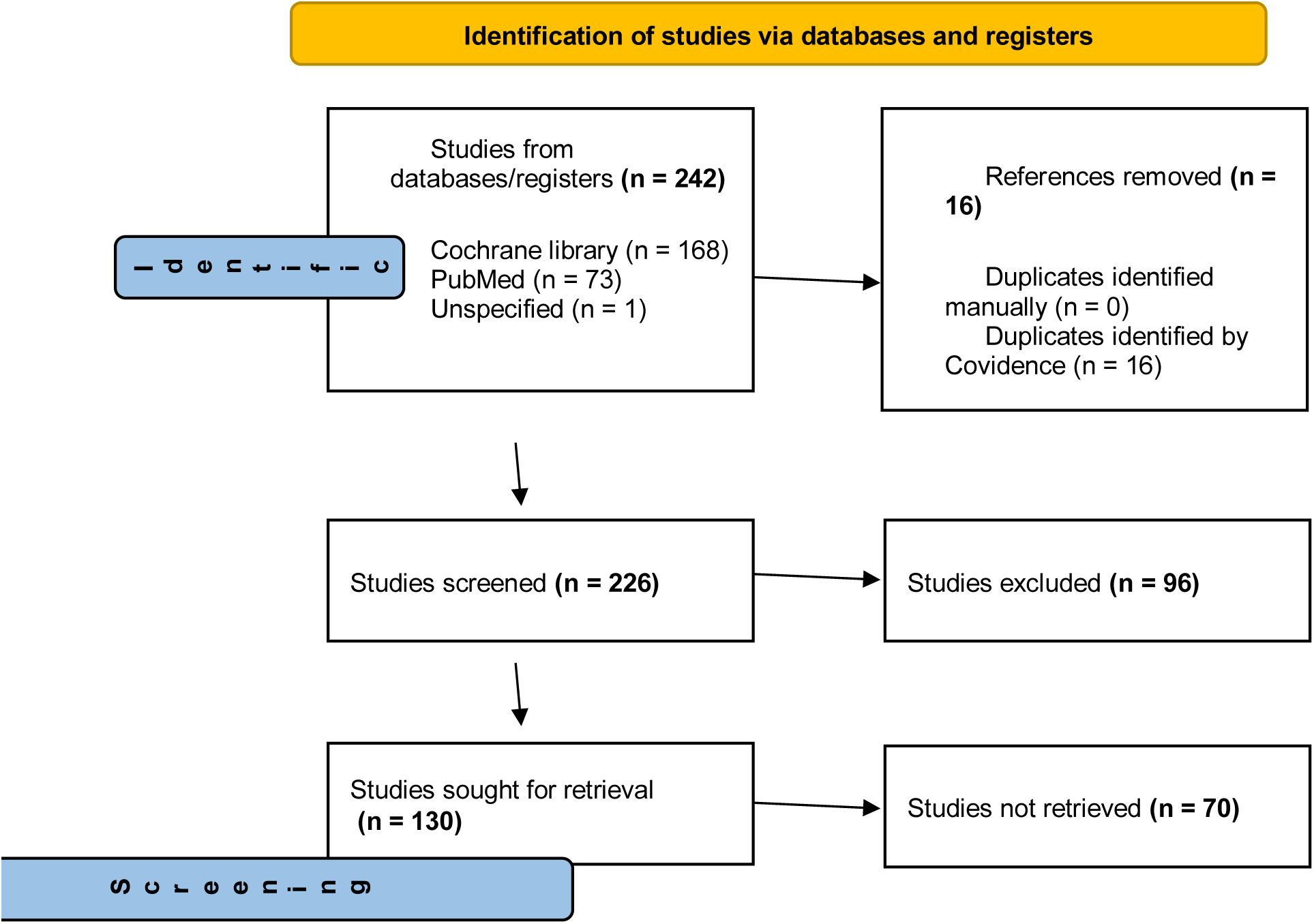
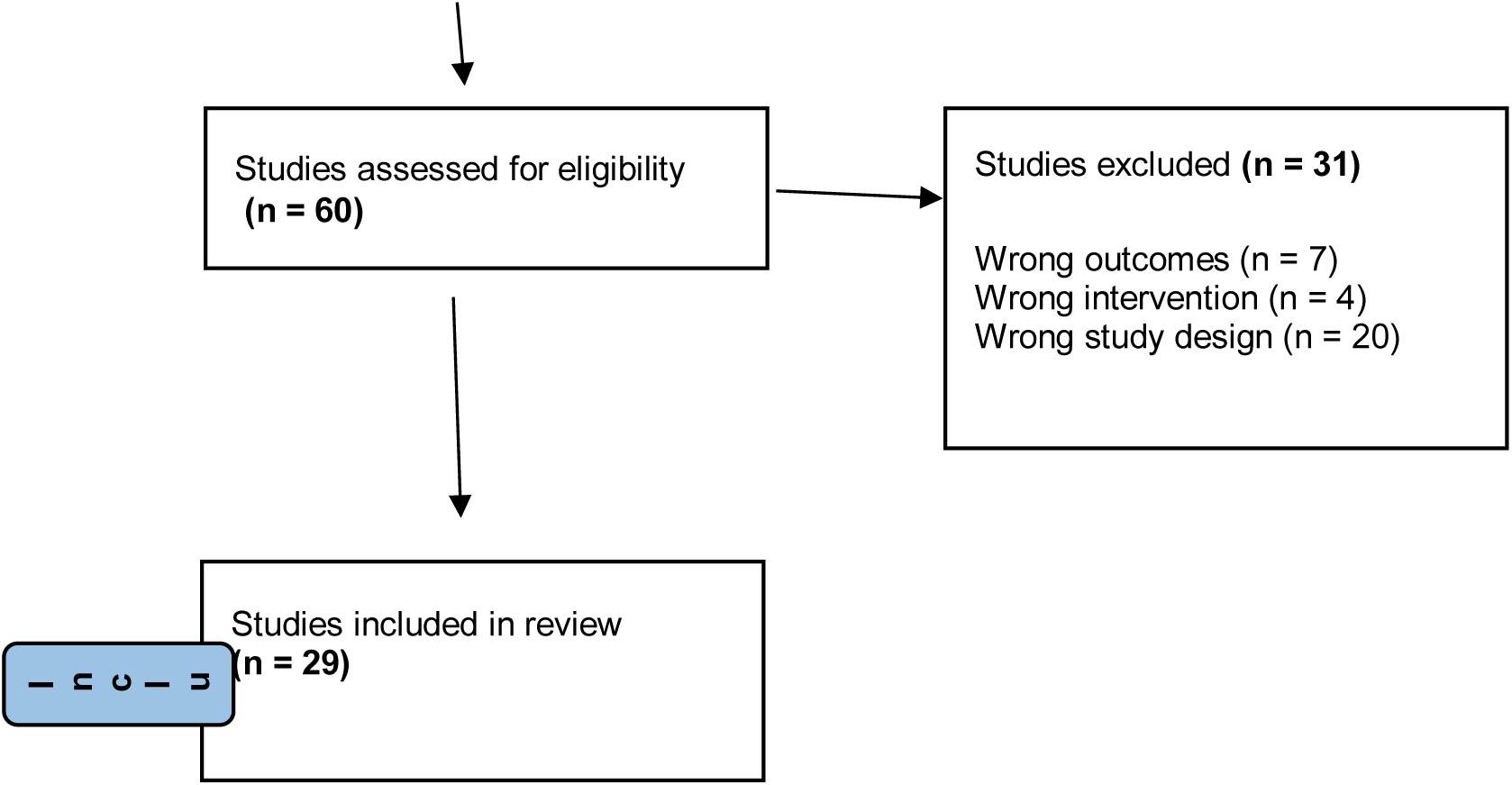
PRISMA flow diagram of the selection process of records

**Fig:2:**
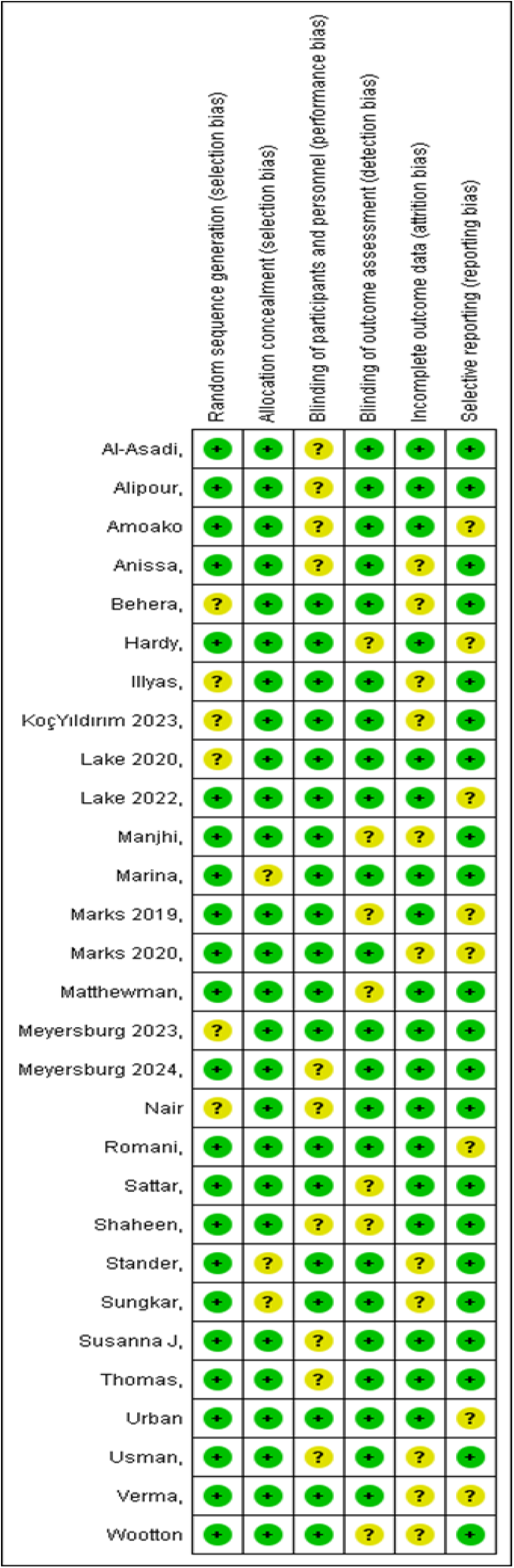
Risk of bias summary: review authors’ judgments about each risk of bias item for each included study.

### 3.2. Risk of Bias of Included Studies

Overall, the studies were rated low to moderate risk of bias. None of the studies were rated as a high risk of bias. All types of bias (Confounding, selection, exposure assessment, outcome measurement, missing data, and selective reporting) were graded as low to moderate only.

**Fig: 1:**
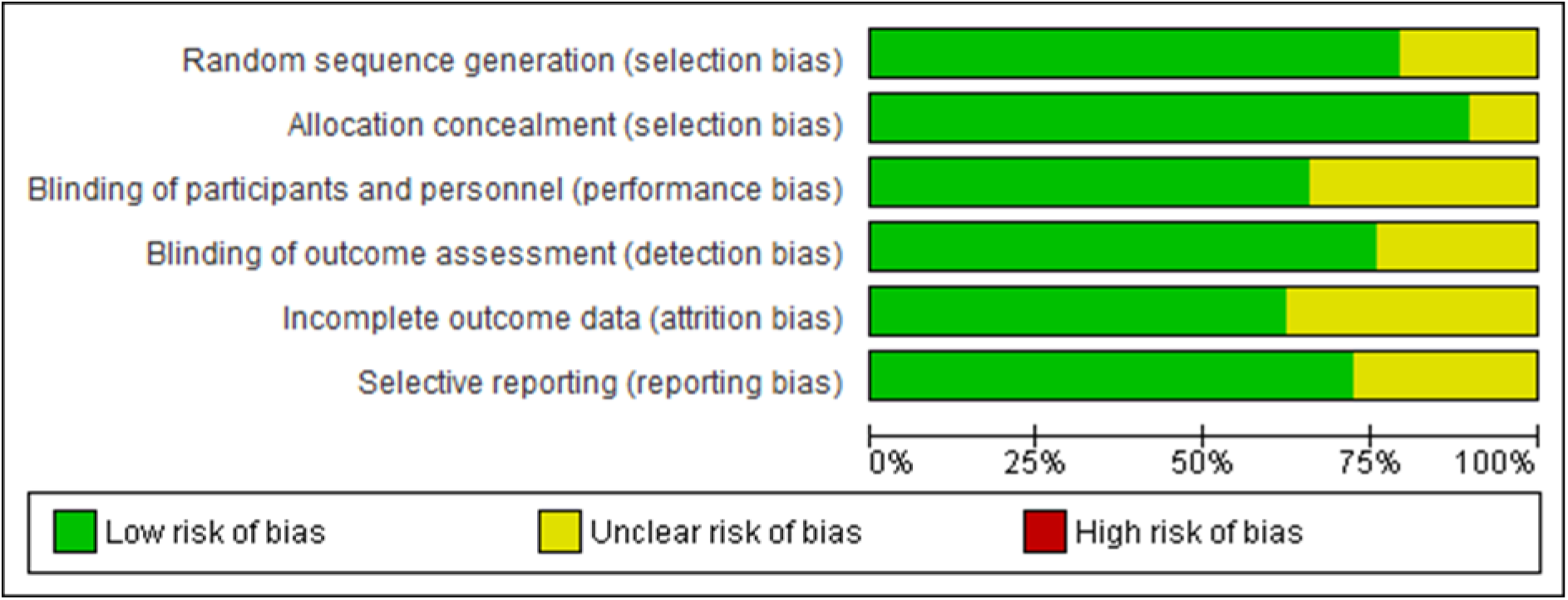
Risk of bias graph: review authors’ judgments about each risk of bias item presented as percentages across all included studies.

### 3.3. General study characteristics

The study involved a significant sample size, comprising data from 40,343 individuals across 20 villages, 6 communities, and 50 countries. This extensive geographic coverage enabled a broad analysis of treatment regimens. The treatment durations studied ranged from 4 weeks to 12 months, reflecting the real-world variability in how long individuals receive treatments. Within this scope, the study design incorporated 22 randomized controlled trials (RCTs) and 2 cross-sectional studies.

Table 1 gives an overview of the 29 studies included in the systematic review. The publication years ranged from 2014 to 2024.

**Table. 1.** Selection Criteria.

|  |  |
| --- | --- |
| Population | Patients with risk factors, physical symptoms, and stigma associated with scabies |
| Intervention | Recent advancements in the management of scabies |
| Comparison | Comparison with previously available treatment options |
| Outcome | Impact on the Quality of life of patients |
| Study Design | RCT and clinical Trials |

### 3.4. Conceptualization, operationalization, rationale, and discussion of results

All 22 studies demonstrated a significant impact of scabies diagnosis and quality of life on the participants. Koç Yıldırım S et al. observed a moderate to severe effect on the quality of life [38], while Khaled Seetan et al. identified a significant association between scabies and anxiety/depression, further compromising the quality of life [39].

In terms of treatment and management, permethrin was found to be the most effective in treating scabies and associated sequelae. It targets eggs, lice, and mites by working on sodium transport across neuronal membranes in arthropods, causing depolarization. This results in the respiratory paralysis of the affected arthropod. Abdel-Raheem et al. noted its high acceptability due to rapid action, although at a higher cost per case. Alternatively, Ivermectin demonstrated efficacy and acceptability rates of 84% and 96%, respectively, at a lower cost compared to permethrin. Benzyl benzoate, though cheaper, exhibited lower efficacy rates. Sulphur ointment, despite being the most expensive option, provided the least favorable outcomes [40]. Certain studies have reported that both permethrin and ivermectin regimens are equally efficacious and well-tolerated in scabies [41].

When comparing permethrin to other alternatives, Abolfazl et al. found that two applications of permethrin 5% cream matched the efficacy of single applications of crotamiton 10% cream at the 2-week follow-up, with superior outcomes for permethrin at the 4-week follow-up. Similarly, Goldust et al. reported significantly higher efficacy with permethrin 2.5% cream compared to Tenutex emulsion at the 4-week follow-up [42].

Given the developing resistance against Permethrin, Alipour et al. mentioned that Ivermectin can also treat scabies effectively. A single dose of ivermectin provided a cure rate of 61.9% at the 2-week follow-up, which increased to 78.5% at the 4-week follow-up after repeating the treatment [43]. The drug binds to glutamate-gated chloride channels common to invertebrate nerve and muscle cells. The binding pushes the channels open, which increases the flow of chloride ions and hyperpolarizes the cell membranes, paralyzing and killing the arthropod. Alternatively, combining drugs also helped to prevent resistance. <u>Michael</u> <u>Marks</u> et al. suggested that co-administration of azithromycin with ivermectin led to similar decreases in scabies prevalence compared to ivermectin [44]. Alebiosu et al. reported that Toto ointment and soap are particularly efficacious in the management of scabies, suggesting that they can be included in the treatment regimen to prevent resistance. [45]

## Discussion

Scabies remains a significant global health issue, particularly in resource-limited settings. Despite its prevalence and impact on individuals and communities, scabies often receive inadequate attention in healthcare settings and research agendas. Scabies affects individuals of all ages and socioeconomic backgrounds, with a higher prevalence observed in overcrowded areas. The burden of scabies extends beyond physical discomfort, impacting quality of life, mental health, and economic productivity. Thus, it is imperative to diagnose scabies early and initiate treatment as soon as possible.

Diagnosis can be made through a thorough elicitation of history and clinical examination. A history of new, intense pruritus and dermatitis raises suspicion for diagnosis, which can be confirmed by skin scrapings and microscopic demonstration of scabies mites, eggs, or fecal matter. [46]

Recent advances in diagnosis include PCR assays to screen for scabies mite DNA. Dermatoscopy, which is a non-invasive imaging technique, has shown promising results in this regard. Other non-invasive procedures, including video dermatoscopy, reflectance confocal microscopy, and optical coherence tomography, have demonstrated improved efficacy in the diagnosis of scabies. [47]

Difficulty in diagnosis can arise in many situations, as scabies present with intense itching and characteristic burrows, but these signs may be absent in certain individuals, leading to misdiagnosis or delayed diagnosis. The similarity of symptoms with other skin conditions like eczema or dermatitis complicates accurate diagnosis, especially in settings where diagnostic tools are limited.

According to the European Academy of Dermatology and Venereology, recommended treatments are Permethrin 5% cream, oral ivermectin, and benzyl benzoate 25% lotion. Alternative treatment modalities are Malathion 0.5% aqueous solution, ivermectin 1% lotion, and sulfur 6-33% cream, ointment, or lotion.

Crusted scabies therapy requires topical scabicide or oral ivermectin. Mass treatments of large populations with endemic disease can be done with a single dose of ivermectin at 200 mg/kg body weight. Any or all sexual partners for the past two months must also be treated.

The STI workup of affected individuals is indicated. Patients and close contacts must abstain from sexual contact until the completion of treatment and should observe strict personal hygiene when living in crowded spaces. [48]

Known causes of treatment failure include improper application of external agents, failure of repeated treatment with ivermectin, incomplete decontamination of belongings, failure to treat people in close contact simultaneously, and other risk factors for the patient. Some documentation suggests a poor response to permethrin due to the resistance of scabies mites. [46]

Recent advancements in the treatment of scabies are explored, including new topical ointments and oral regimens. There are trials to bring in new drugs to avoid resistance to drug therapy. Moxidectin is a new drug that is undergoing clinical trials to replace permethrin in regions with high resistance. [46]

Challenges in treatment include resistance to conventional chemotherapy due to overreliance on standard treatments like permethrin or ivermectin, reducing treatment efficacy. Treating scabies in certain demographics, such as infants, pregnant women, or immunocompromised individuals, requires careful consideration due to safety concerns associated with conventional therapies.

Since scabies is considered a significant health problem worldwide, intensive research looks into aspects of the biology of Sarcoptes scabiei and host-parasite interactions. However, the practicality of conducting these trials is limited due to the inability to maintain mites in vitro and the limited access to infective material. At present, research is focused on the initial characterization of molecules of interest to improve diagnosis and vaccines and eliminate drug resistance. Recent developments anticipate combining animal models and next-generation technologies to create new strategies to prevent, diagnose, and treat scabies, ultimately improving skin health in patients. [49]

The incorporation of telemedicine and ethical AI in medicine can also improve the diagnosis and early treatment of scabies in patients. Exploration of host factors influencing scabies susceptibility and treatment response to develop personalized treatment approaches. Utilization of machine learning algorithms for early detection of scabies outbreaks, predicting treatment responses, and identifying genetic markers associated with scabies resistance can be areas of potential research in the future.

To improve patient outcomes in scabies, certain comprehensive strategies, like early diagnosis and treatment compliance, should be followed. A prompt diagnosis of scabies can cut down on transmission and prevent further complications. Consistent treatment ensures the complete eradication of the disease and prevents resistance. Educating patients about proper hygiene, transmission, treatment compliance, and prevention of scabies plays a vital role in reducing the global burden of scabies. Healthcare individuals must be equipped with the latest diagnostic technologies and knowledge of the latest treatment guidelines to improve patient outcomes. Resistant cases must be addressed at the earliest possible time to prevent further transmission. Finally, effective research focusing on developing newer diagnostic and treatment modalities must be conducted for improved patient outcomes.

## Conclusion

Our study concluded that scabies is a debilitating disease with a significant impact on quality of life. It was found that permethrin was the most effective treatment modality and was widely accepted by all patients. It also has a rapid onset of action; however, the higher cost per patient may limit its use in every region. While Ivermectin is a drug that is better used in cases of permethrin-resistant scabies, it has a lower cost and can be used as a first-line drug. Among all the drugs, benzyl benzoate is the cheapest, and sulfur ointments are the most expensive. The combination of drugs has yielded significant positive results and should be explored more. Recent advances include efforts to minimize drug resistance and increase the efficacy of existing drugs. With drugs like moxidectin undergoing trials for usage in areas of high resistance [46] and other drugs for topical use, hope for the elimination of this disease ensues.

With studies describing the impairment of QoL as being ’ very large’ [50], the need to address the symptoms and provide complete and effective care is rising. There are multiple studies on the quality of life, but these are very scattered and region-specific. A global study must be done to evaluate children and adults from all over the world, and interventions should be implemented at a grass-roots level. The effect of these interventions should be studied concerning improvement in QoL. Interventions should be both ‘patient-centered’ to allow a rapid cure of disease with minimal psychosocial effect in the person’s daily life. Drugs can be developed that are both anti-parasite and prevent secondary bacterial infections.

Scabies was declared a neglected tropical disease [NTD] in 2017 by the WHO [51]. Since then, multiple programs have been set forth to target the reduction of the prevalence of these NTDs and promote access to rapid-acting and cost-effective medications to treat this disease. The World Scabies Program [52] is another worldwide project that aims to spread awareness about the health implications of this disease and promote groundbreaking translational research to enhance the development of drugs and treatment strategies. An urgent global collaboration is key to the control and management of this disease. Surveillance and interventional programs must be conducted at every level of the community, starting from the smallest villages and rural areas to the state and country as a whole. Control requires an interdisciplinary approach, involving doctors, community workers, state administration, psychologists, and epidemiologists, among other stakeholders.

## Data Availability

All data produced in the present study are available upon reasonable request to the authors

## References

1. Salavastru CM, Chosidow O, Boffa MJ, Janier M, Tiplica GS. European guideline for the management of scabies. J Eur Acad Dermatol Venereol. 2017 Aug;31(8):1248–53.

2. Micali G, Lacarrubba F, Verzì AE, Chosidow O, Schwartz RA. Scabies: Advances in Noninvasive Diagnosis. Vinetz JM, editor. PLoS Negl Trop Dis. 2016 Jun 16;10(6):e0004691.

3. Paudel S, Pudasaini P, Adhikari S, Pradhan MB, Shekhar Babu KC. Quality of life in patients with scabies: A cross-sectional study using Dermatology Life Quality Index (DLQI) questionnaire. JEADV Clin Pract. 2023 Jun;2(2):399–403.

4. Koç Yıldırım S, Demirel Öğüt N, Erbağcı E, Öğüt Ç. Scabies Affects Quality of Life in Correlation with Depression and Anxiety. Dermatol Pract Concept. 2023 Apr 29;e2023144.

5. Marks M, Toloka H, Baker C, Kositz C, Asugeni J, Puiahi E, et al. Randomized Trial of Community Treatment With Azithromycin and Ivermectin Mass Drug Administration for Control of Scabies and Impetigo. Clin Infect Dis. 2019 Mar 5;68(6):927–33.

6. Alipour H, Goldust M. The efficacy of oral ivermectin vs. sulfur 10% ointment for the treatment of scabies. Ann Parasitol. 2015;61(2):79–84.

7. Goldust M, Rezaee E, Raghifar R, Hemayat S. Treatment of scabies: the topical ivermectin vs. permethrin 2.5% cream. Ann Parasitol. 2013;59(2):79– 84.

8. Abdel-Raheem TA, Méabed EMH, Nasef GA, Abdel Wahed WY, Rohaim RMA. Efficacy, acceptability and cost effectiveness of four therapeutic agents for treatment of scabies. J Dermatol Treat. 2016 Sep 2;27(5):473–9.

9. Meyersburg D, Hoellwerth M, Brandlmaier M, Handisurya A, Kaiser A, Prodinger C, et al. Comparison of topical permethrin 5% vs. benzyl benzoate 25% treatment in scabies: a double-blinded randomized controlled trial. 2024;190(4):486–491-. Available from:

10. Hardy M, Samuela J, Kama M, Tuicakau M, Romani L, Whitfeld MJ, et al. Community control strategies for scabies: a cluster randomised noninferiority trial. 2021;18(11):e1003849-. Available from:

11. Alipour H, Goldust M. The efficacy of oral ivermectin vs. sulfur 10% ointment for the treatment of scabies. 2015;61(2):79–84-. Available from:

12. Marina A, Menaldi SL, Novianto E, Widaty S. Efficacy of 5% permethrin-2% fusidic acid cream compared to 5% permethrin-placebo in the treatment of impetiginized scabies. 2022;16(6):1045–1054-. Available from:

13. Romani L, Marks M, Sokana O, Nasi T, Kamoriki B, Cordell B, et al. Efficacy of mass drug administration with ivermectin for control of scabies and impetigo, with coadministration of azithromycin: a single-arm community intervention trial. 2019;19(5):510–518-. Available from:

14. Illyas M, Shahzad A, Siddiqui S, Awais M, Qayyum M, Azfar NA. Comparison of efficacy of oral ivermectin and topical permethrin 5% in the treatment of scabies. 2023;33(1):123–131-. Available from:

15. ACTRN12618001086257. Regimens of Ivermectin for Scabies Elimination: a cluster randomised non-inferiority trial of one versus two doses of ivermectin for the control of scabies using a mass drug administration strategy. 2018; Available from:

16. Matthewman J, Manego RZ, Dimessa Mbadinga LB, Šinkovec H, Völker K, Akinosho M, et al. A randomized controlled trial comparing the effectiveness of individual versus household treatment for Scabies in Lambaréné, Gabon. 2020;14(6):e0008423-. Available from:

17. Lake SJ, Phelan SL, Engelman D, Sokana O, Nasi T, Boara D, et al. Protocol for a cluster-randomised non-inferiority trial of one versus two doses of ivermectin for the control of scabies using a mass drug administration strategy (the RISE study). 2020;10(8):e037305-. Available from:

18. Sungkar S, Agustin T, Menaldi SL, Fuady A, Herqutanto, Angkasa H, et al. Effectiveness of permethrin standard and modified methods in scabies treatment. 2014;23(2):93–98-.

19. Meyersburg D, Welponer T, Kaiser A, Selhofer S, Tatarski R, Handisurya A, et al. Comparison of topical benzyl benzoate vs. oral ivermectin in treating scabies: a randomized study. 2023;37(1):160–165-.

20. Behera P, Munshi H, Kalkonde Y, Deshmukh M, Bang A. Control of scabies in a tribal community using mass screening and treatment with oral ivermectin -A cluster randomized controlled trial in Gadchiroli, India. 2021;15(4):e0009330-.

21. Marks M, Gwyn S, Toloka H, Kositz C, Asugeni J, Asugeni R, et al. Impact of Community Treatment with Ivermectin for the Control of Scabies on the Prevalence of Antibodies to Strongyloides stercoralis in Children. 2020;71(12):3226–3228-.

22. Marks M, Toloka H, Baker C, Kositz C, Asugeni J, Puiahi E, et al. Randomized Trial of Community Treatment With Azithromycin and Ivermectin Mass Drug Administration for Control of Scabies and Impetigo. 2019;68(6):927–933-.

23. Thomas J, Davey R, Peterson GM, Carson C, Walton SF, Spelman T, et al. Treatment of scabies using a tea tree oil-based gel formulation in Australian Aboriginal children: protocol for a randomised controlled trial. 2018;8(5):e018507-.

24. Anissa L, Indriatmi W, Wibawa LP, Widaty S. Efficacy and side effects of Blacksoap® as adjuvant therapy of scabies: a randomized control trial. 2022;31(2):102–107-.

25. Usman A, Shehzad A, Shahid M, Saleem Z, Khawaja A, Abeer. Comparison between efficacy of topical 1% ivermectin and topical 5% permethrin in the treatment of scabies. 2021;31(4):633–636-.

26. Shaheen A, Usman S, Mirbahar AM, Hameed RMH. Comparing the two treatment regimes for scabies, topical permethrin 5% with oral ivermectin. 2017;28(3):15–18-. Available from:

27. Al-Asadi ZA, Al-Hamdi KI, Ahmed JH. Effectiveness of Combined Oral and Topical Ivermectin Compared to Topical Treatments in Patients with Scabies. 2023;13(1):101–104-. Available from:

28. Manjhi PK, Sinha RI, Kumar M, Sinha KI. Comparative study of efficacy of oral ivermectin versus some topical antiscabies drugs in the treatment of scabies. 2014;8(9):HC01–4-.

29. Sattar RMT, Malik NA, Riaz U, Khalid AA, Jabin I, Warraich FK. Comparison of Efficacy of Single Application Topical 5% Permethrin versus Single Dose Oral Ivermectin in the Treatment of Scabies. 2023;17(10):18–20-.

30. Verma S, Ahsan M. Comparison of topical ivermectin and oral ivermectin in the treatment of human scabies: a randomized controlled trial. 2020;11(4):134–139

31. Lake SJ, Engelman D, Sokana O, Nasi T, Boara D, Marks M, et al. Health-related quality of life impact of scabies in the Solomon Islands. 2022;116(2):148–156-.

32. Ständer S. Itch in Scabies-What Do We Know? Front Med (Lausanne). 2021;8:628392.

33. Koç Yıldırım S, Demirel Öğüt N, Erbağcı E, Öğüt Ç. Scabies Affects Quality of Life in Correlation with Depression and Anxiety. Dermatol Pract Concept. Published online April 1, 2023. doi:10.5826/dpc.1302a144

34. Amoako YA, S van RL, Oppong MN, Amoako KO, Abass KM, Anim BA, et al. Beliefs, attitudes and practices towards scabies in central Ghana. PLoS Negl Trop Dis. 2023;17(2):e0011175-.

35. Wootton CI, Bell S, Philavanh A, Phommachack K, Soukavong M, Kidoikhammouan S, et al. Assessing skin disease and associated health-related quality of life in a rural Lao community. BMC Dermatol. 2018;18(1):11.

36. Nair PA, Vora R V, Jivani NB, Gandhi SS. A Study of Clinical Profile and Quality of Life in Patients with Scabies at a Rural Tertiary Care Centre. J Clin Diagn Res. 2016;10(10):WC01–5.

37. Urban K, Chu S, Giesey RL, Mehrmal S, Uppal P, Delost ME, et al. Burden of skin disease and associated socioeconomic status in Asia: A cross-sectional analysis from the Global Burden of Disease Study 1990-2017. JAAD Int. 2021;2:40–50.

38. Koç Yıldırım S, Demirel Öğüt N, Erbağcı E, Öğüt Ç. Scabies Affects Quality of Life in Correlation with Depression and Anxiety. Dermatol Pract Concept. 2023 Apr 1;13(2):e2023144. doi: 10.5826/dpc.1302a144. Epub ahead of print. PMID: 37196304; PMCID: PMC10188156.

39. Seetan, Khaled, Balqis Abusohyon Almu’atasim Khamees, Hedaia Mohammad Alrababah, Furat Khasawneh, Ahmad Mowafaq Albataineh, Ali Zaid Mahajnah, and Rahaf Al-Showaiter. “SCABIES IS ASSOCIATED WITH MORE ANXIETY, DEPRESSION, AND IMPAIRED QUALITY OF LIFE: A CROSS-SECTIONAL COMPARATIVE STUDY.” Journal of Pharmaceutical Negative Results (2022): 6314–6320.

40. Abdel-Raheem TA, Méabed EM, Nasef GA, Abdel Wahed WY, Rohaim RM. Efficacy, acceptability and cost effectiveness of four therapeutic agents for treatment of scabies. J Dermatolog Treat. 2016 Oct;27(5):473–9. doi: 10.3109/09546634.2016.1151855. Epub 2016 Mar 30. PMID: 27027929.

41. Ranjkesh MR, Naghili B, Goldust M, Rezaee E. The efficacy of permethrin 5% vs. oral ivermectin for the treatment of scabies. Ann Parasitol. 2013;59(4):189–94. PMID: 24791346.

42. Goldust M, Rezaee E, Raghifar R, Hemayat S. Treatment of scabies: the topical ivermectin vs. permethrin 2.5% cream. Ann Parasitol. 2013;59(2):79–84. PMID: 24171301.

43. Alipour H, Goldust M. The efficacy of oral ivermectin vs. sulfur 10% ointment for the treatment of scabies. Ann Parasitol. 2015;61(2):79–84. PMID: 26342502.

44. Marks M, Toloka H, Baker C, Kositz C, Asugeni J, Puiahi E, Asugeni R, Azzopardi K, Diau J, Kaldor JM, Romani L, Redman-MacLaren M, MacLaren D, Solomon AW, Mabey DCW, Steer AC. Randomized Trial of Community Treatment With Azithromycin and Ivermectin Mass Drug Administration for Control of Scabies and Impetigo. Clin Infect Dis. 2019 Mar 5;68(6):927–933. doi: 10.1093/cid/ciy574. PMID: 29985978; PMCID: PMC6399435.

45. Alebiosu CO, Ogunledun A, Ogunleye DS. A report of clinical trial conducted on Toto ointment and soap products. J Natl Med Assoc. 2003 Jan;95(1):95–105. PMID: 12656456; PMCID: PMC2594367.

46. Salavastru CM, Chosidow O, Boffa MJ, Janier M, Tiplica GS. European guideline for the management of scabies. J Eur Acad Dermatol Venereol. 2017 Aug;31(8):1248–1253. doi: 10.1111/jdv.14351. Epub 2017 Jun 22. PMID: 28639722

47. Sunderkötter C, Wohlrab J, Hamm H. Scabies: Epidemiology, Diagnosis, and Treatment. Dtsch Arztebl Int. 2021 Oct 15;118(41):695–704. doi: 10.3238/arztebl.m2021.0296. PMID: 34615594; PMCID: PMC8743988.

48. Micali G, Lacarrubba F, Verzì AE, Chosidow O, Schwartz RA. Scabies: Advances in Noninvasive Diagnosis. PLoS Negl Trop Dis. 2016 Jun 16;10(6):e0004691. doi: 10.1371/journal.pntd.0004691. PMID: 27311065; PMCID: PMC4911127.

49. Mounsey KE, McCarthy JS, Walton SF. Scratching the itch: new tools to advance understanding of scabies. Trends Parasitol. 2013 Jan;29(1):35–42. doi: 10.1016/j.pt.2012.09.006. Epub 2012 Oct 19. PMID: 23088958.

50. Paudel S, Pudasaini P, Adhikari S, Pradhan M, Babu KCS. Quality of life in patients with scabies: A cross-sectional study using Dermatology Life Quality Index (DLQI) questionnaire. JEADV Clinical Practice. 2023;2(2):399–403. doi:10.1002/jvc2.127

51. World Health Organization: WHO, World Health Organization: WHO. Scabies. Published May 31, 2023. https://www.who.int/news-room/fact-sheets/detail/scabies

52. Home - World Scabies program. https://www.worldscabiesprogram.org/

